# Machine learning identified distinct serum lipidomic signatures in hospitalized COVID-19-positive and COVID-19-negative patients

**DOI:** 10.1101/2021.12.14.21267764

**Authors:** Helena Castañé, Simona Iftimie, Gerard Baiges-Gaya, Elisabet Rodríguez-Tomàs, Andrea Jiménez-Franco, Ana Felisa López-Azcona, Pedro Garrido, Antoni Castro, Jordi Camps, Jorge Joven

**Affiliations:** Unitat de Recerca Biomèdica, Hospital Universitari de Sant Joan, Institut d’Investigació Sanitària Pere Virgili, Universitat Rovira i Virgili, Reus, Spain; Department of Internal Medicine, Hospital Universitari de Sant Joan, Institut d’Investigació Sanitària Pere Virgili, Universitat Rovira i Virgili, Reus, Spain; Intensive Care Unit, Hospital Universitari de Sant Joan, Institut d’Investigació Sanitària Pere Virgili, Universitat Rovira i Virgili, Reus, Spain

**Author notes:** Corresponding author at: Unitat de Recerca Biomèdica, Hospital Universitari de Sant Joan, C. Sant Joan s/n, 43201 Reus, Spain., *E-mail address* (J. Camps). Helena Castañé and Simona Iftimie contributed equally to this manuscript.

**Keywords:** artificial intelligence, COVID-19, lipid metabolism, lipidomics, machine learning

## Abstract

**Background:** Lipids are involved in the interaction between viral infection and the host metabolic and immunological response. Several studies comparing the lipidome of COVID-19-positive hospitalized patients *vs*. healthy subjects have already been reported. It is largely unknown, however, whether these differences are specific to this disease. The present study compared the lipidomic signature of hospitalized COVID-19-positive patients with that of healthy subjects, and with COVID-19-negative patients hospitalized for other infectious/inflammatory diseases. Potential COVID-19 biomarkers were identified.

**Methods:** We analyzed the lipidomic signature of 126 COVID-19-positive patients, 45 COVID-19-negative patients hospitalized with other infectious/inflammatory diseases and 50 healthy volunteers. Results were interpreted by machine learning.

**Results:** We identified acylcarnitines, lysophosphatidylethanolamines, arachidonic acid and oxylipins as the most altered species in COVID-19-positive patients compared to healthy volunteers. However, we found similar alterations in COVID-19-negative patients. By contrast, we identified lysophosphatidylcholine 22:6-sn2, phosphatidylcholine 36:1 and secondary bile acids as the parameters that had the greatest capacity to discriminate between COVID-19-positive and COVID-19-negative patients.

**Conclusion:** This study shows that COVID-19 infection shares many lipid alterations with other infectious/inflammatory diseases, but differentiating them from the healthy population. Also, we identified some lipid species the alterations of which distinguish COVID-19-positive from Covid-19-negative patients. Our results highlight the value of integrating lipidomics with machine learning algorithms to explore the pathophysiology of COVID-19 and, consequently, improve clinical decision making.

## 1. Introduction

COVID-19 infection produces dramatic changes in the metabolism of the infected cells, including the concentration and composition of different lipid species [1,2]. Lipids combine with thousands of metabolites and hundreds of specific pathways in support of the life-cycle of the organism [3]. As such, it is not surprising that lipids are involved in the interplay between viral infection and the host’s response [2]. Viruses enter cells through protein-lipid interactions [4,5], are externalized via lipid vesicles [6], while interactions between viruses and the organism alter mitochondrial metabolism and the microbiota [7-9] that have important implications in body lipid composition. Further, oxidative stress triggered by infection results in oxidized lipids production by specific biosynthetic pathway activation [10]. Studies comparing the lipidome of COVID-19-positive patients *vs*. healthy subjects have been reported and distinctive lipid species have been identified [2]. However, there is a paucity of information regarding the specificity of these measurements, i.e. whether variations in circulating levels of the species identified are characteristic of the COVID-19 infection, or whether they can be observed in other infectious or inflammatory diseases, as well. Our study was aimed at identifying alterations in the serum lipidome of patients with COVID-19 infection, the aim being to evaluate the relationships between the alterations and the disease with a view to identifying potential biomarkers that would help in clinical decisions in diagnosis and treatment.

## 2. Materials and methods

### 2.1. Study design and participants

We performed a retrospective *post-hoc* cohort study in 126 patients hospitalized for COVID-19 between March and October 2020 in the Department of Internal Medicine, or in the Intensive Care Unit (ICU) of our Institution. Inclusion criteria into the present study were: ≥18 years of age and a positive PCR result for COVID-19 obtained within 24 hours before the blood sample was drawn for the study. Exclusion criteria were: having a life expectancy ≤24 hours, impaired liver function, or pregnancy. We also analyzed samples from 45 COVID-19-negative patients hospitalized with diseases having an infectious/inflammatory component. These samples, collected in 2019, belonged to a previous prospective study in patients with urinary catheter-related infection. A detailed description of these patients has been published [11]. For the purposes of the present study, we selected a subgroup with a distribution of age and sex to match, as closely as possible, the COVID-19-positive patients. As a control group, we analyzed samples from 50 healthy volunteers who had participated in an epidemiological study, the details of which have already been reported [12]. The subjects had no clinical or biochemical evidence of diabetes, cancer, kidney failure, liver disease, or neurological disorders. Serum samples from all participants were stored in our Biobank at –80ºC until the time of batched analyses. We recorded clinical and demographic data and calculated the McCabe score as an index of clinical prognosis [13] and the Charlson index as a way of categorizing patient comorbidities [14]. This study was approved by the *Comitè d’Ètica i Investigació en Medicaments* (Institutional Review Committee) of the *Institut d’Investigació Sanitària Pere Virgili* (Resolution CEIM 040/2018, modified on April 16, 2020).

### 2.2. Lipidomics analyses

Lipids were analyzed by semi-targeted lipidomics. Methods have been previously reported by our research group [15,16] and are described in detail in Supplementary Methods 1 (Supplementary_Material.docx file). Briefly, acylcarnitines (CAR) were extracted with methanol and analyzed by triple quadrupole liquid chromatography/mass spectrometry (LC-QqQ). Non-polar lipids were extracted with a mixture of tert-butyl ether and methanol (1:2 v/v) with 0.5% acetic acid and analyzed by quadrupole time-of-flight liquid chromatography/mass spectrometry (LC-qTOF). Polar lipids were extracted with methanol and analyzed by LC-qTOF. Lipids were then matched with the Metlin database (Scripps Research Institute, La Jolla, CA,) and quantified with calibration curves generated with internal standards.

### 2.3. Statistical analyses

Statistical assessments were performed with the R program (RStudio version 4.0.5). The MetaboAnalystR package was used to generate scores and loading plots and included False Discovery Rates (FDR), Volcano plots, Principal Component Analysis (PCA), Partial Least Square Discriminant Analysis (PLS-DA), and hierarchically clustered heatmaps [17]. To evaluate the diagnostic accuracy of different combinations of lipids, we constructed a Monte Carlo cross validation model that combined from 5 to 100 random variables, and subsequently calculated the area under the curve of the Receiver Operating Characteristics (ROC) curves, and confusion matrices [18]. The TableOne package was used to generate mean and standard deviation of all lipid concentrations [19]. The R-commands employed are shown as Supplementary Methods 2 (Supplementary_Materials.docx file).

## 3. Results

### 3.1. Clinical characteristics of the studied groups

The clinical characteristics of all participants are shown in Table 1. COVID-19-negative patients were significantly older and consumed less alcohol than the control group. COVID-19-positive patients had a lower frequency of smoking habit, alcohol intake, type 2 diabetes mellitus, chronic kidney disease and cancer than COVID-19-negative patients. The McCabe score and the Charlson index indicated that COVID-19-positive patients were, in general, less severe than COVID-19-negative patients.

**Table 1.**
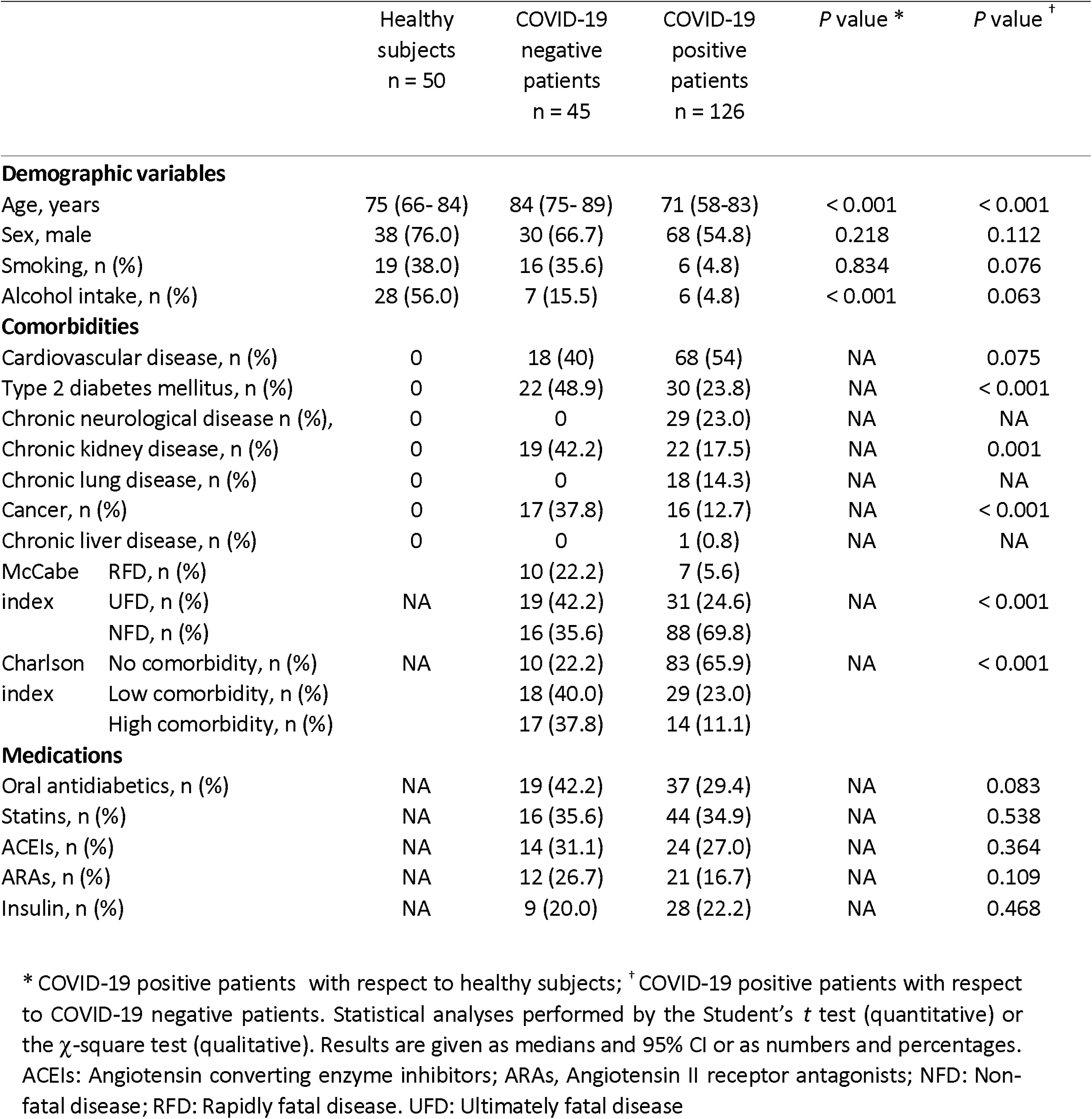
Demographic and clinical characteristics of the patients and the healthy subjects

### 3.2. Acylcarnitines, arachidonic acid and oxylipins: The common lipid signature of COVID-19-positive and COVID-19-negative patients

A total of 283 lipid species were analyzed. Numerical results are shown in Supplementary Tables (Supplementary_Tables.xls file). Volcano plots identified changes in the concentrations of 107 species comparing the COVID-19-positive patients vs. the healthy volunteers and 108 species comparing the COVID-19-negative patients vs. the healthy volunteers. The species with the greatest changes were O-octanoyl-R-carnitine (CAR 8.0) and lysophosphatidylethanolamines (LPE), which were increased, and the oxylipins 9/13-hydroxyoctadecadienoic acid (9-HODE/13-HODE) and 15-hydroxyeicosatetraenoic acid (15-HETE) which were decreased (Fig. 1A). The heatmap clustering algorithm grouped the lipids into four blocks: The first three blocks were constituted mainly by oxylipins and the fourth by bile acids (Fig. 1B). PCA and PLS-DA completely segregated the populations of healthy volunteers from COVID-19 patients (either positive or negative), and the Variable Importance in Projection (VIP) score identified 9-HODE/13-HODE and 15-HETE as the most effective lipids in distinguishing the groups of patients from the healthy volunteers (Fig. 1C and D). We did not observe any significant difference between the position of the fatty acid chain of the lysophospholipids (lysophosphatidylcholines and LPE) in the different study groups (Supplementary Fig. 1).

**Fig. 1.**
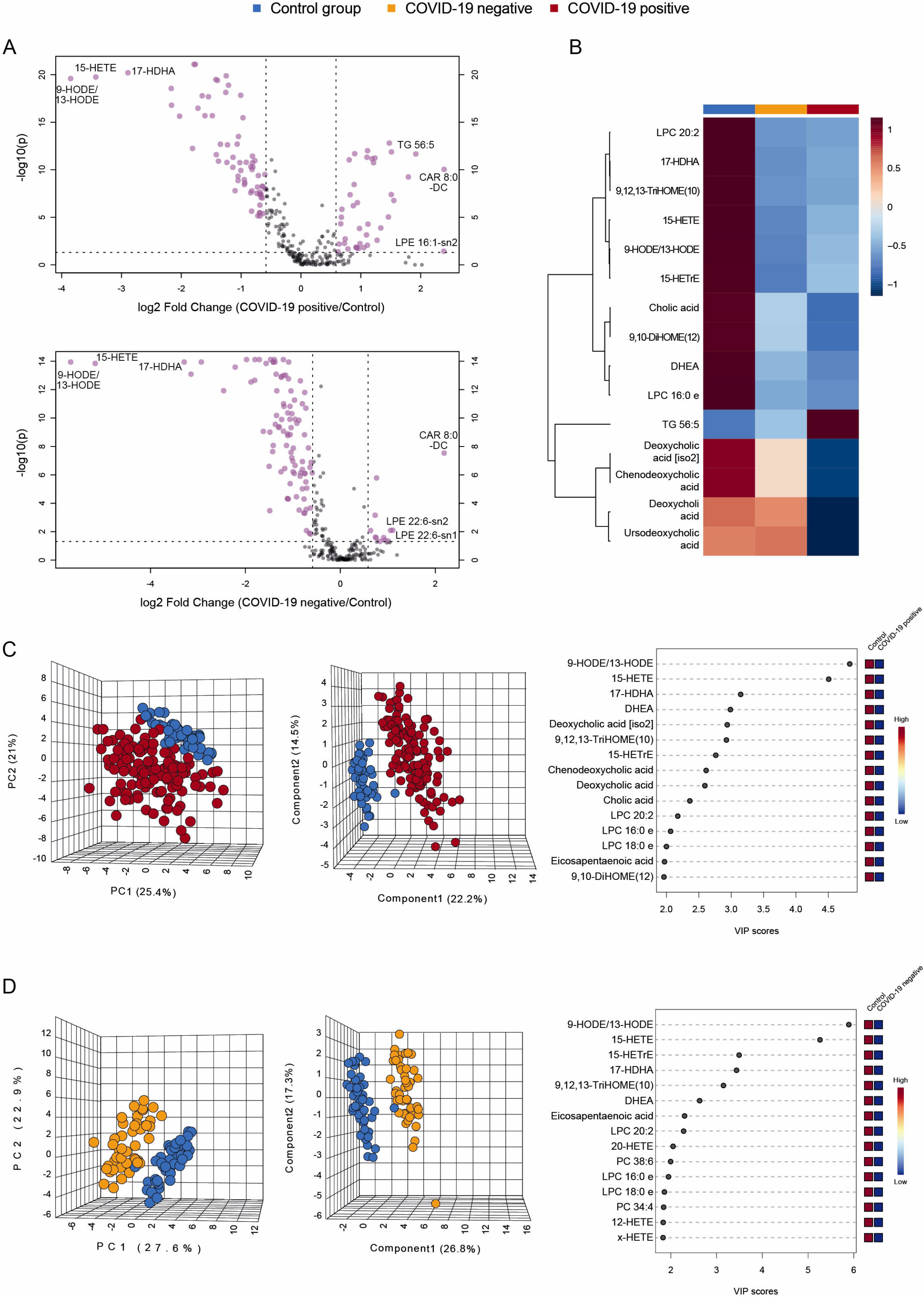
Lipid signatures differentiate COVID-19 positive and COVID-19-negative patients from healthy individuals. (A): Volcano plots representing the log fold-change of lipid species in COVID-19-positive (upper panel) and COVID-19-negative (lower panel) patients relative to the control group. (B): Heatmap showing the 15 most relevant lipid species in the control group (blue), COVID-19-negative (yellow) and COVID-19-positive (red) patients. (C): From left to right: Principal Component Analysis (PCA) clustering of the COVID-19-positive patients and the control group; Principal Least Square Discriminant Analysis (PLS-DA) clustering the COVID-19-positive patients and the control group; Variable Importance in Projection (VIP) score identifying 9/13-HODE and 15-HETE as the most relevant parameters discriminating between COVID-19-positive patients and the control group. (D): From left to right: PCA clustering the COVID-19-negative patients and the control group; PLS-DA clustering the COVID-19-negative patients and the control group; VIP score identifying 9/13-HODE and 15-HETE as the most relevant parameters discriminating between COVID-19-negative patients and the control group. In 3-dimensional plots of PCA and PLS-DA, each ball represents a patient, and positions depend on differences in lipid concentrations. Axes are formed by different combinations of variables, and the percentages represent the proportion of variance that can be explained. PCA is a non-supervised test and PLS-DA is a supervised analysis. Acronyms: CAR: Acylcarnitine; DHEA: dehydroepiandrosterone; DHOME: dihydroxyoctadecenoic acid; HDHA: hydroxydocosahaxaenoic aid; HETE: hydroxyeicosatetraenoic acid; HODE: hydroxyoctadecadienoic acid; LPC: lysophosphatidylcholine; LPE: lysophosphatidylethanolamine; TG: triglyceride.

The enrichment analysis showed an alteration of the pathways of fatty acid synthesis, the metabolism of arachidonic, linoleic and linolenic acids (precursors of oxylipins), and the β-oxidation of fatty acids in COVID-19-positive or COVID-19-negative patients compared to control subjects (Fig. 2A and B).

**Fig. 2.**
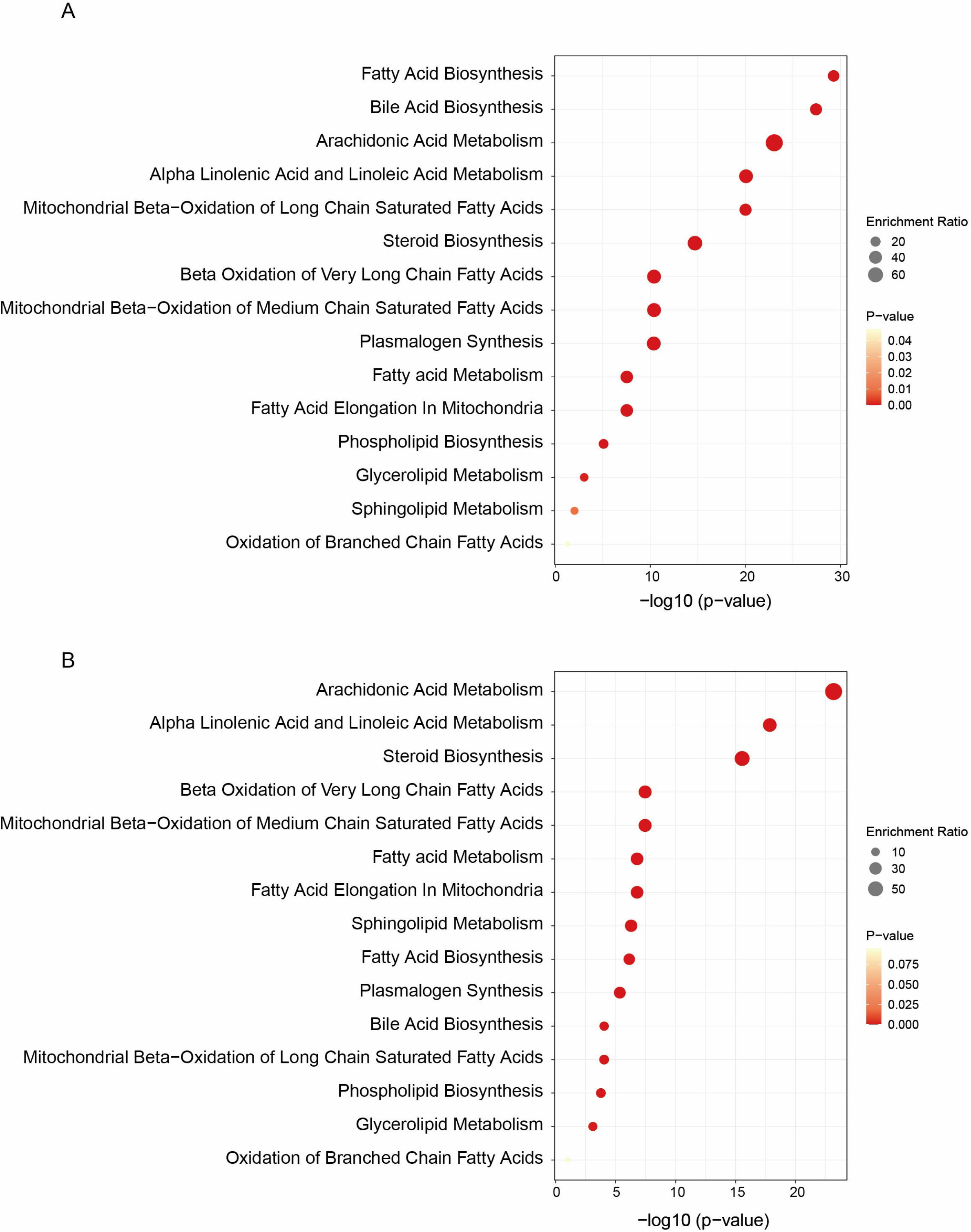
Enrichment analysis showing the most severely affected biochemical pathways in COVID-19-positive patients (A) and COVID-19-negative patients (B) compared with the control group.

Monte Carlo models were generated to help identify those species that could be useful as biomarkers of infectious/inflammatory processes (whether COVID-19-positive or not). The models initially combined five randomly chosen lipid species and determined the area under the curve (AUC) for each from the combined ROC curve. The numbers of variables were progressively increased to 100 in 6 different models. In all instances, the analyses of the AUC-ROC curves were >0.98 (Fig. 3A). The 100-variable model was chosen to construct a confusion matrix, which correctly classified all but 1 of the patients (Fig. 3B). The algorithm identified oxylipins as the most relevant variables in the construction of the model. Other species identified were carnitines and lysophospholipids (Fig. 3C). From among these species, we chose arachidonic acid for further analysis because: its physiological and pathological importance is high; it is one of the main precursors of oxylipin synthesis; its internal standard is commercially available so its quantification is facilitated. Figure 3D shows serum concentrations of arachidonic acid being significantly decreased in COVID-19-positive as well as COVID-19-negative patients. Moreover, the AUCs of the ROC curves for arachidonic acid were >0.97 in the discrimination of both patient groups from the control group (Fig. 3E).

**Fig. 3.**
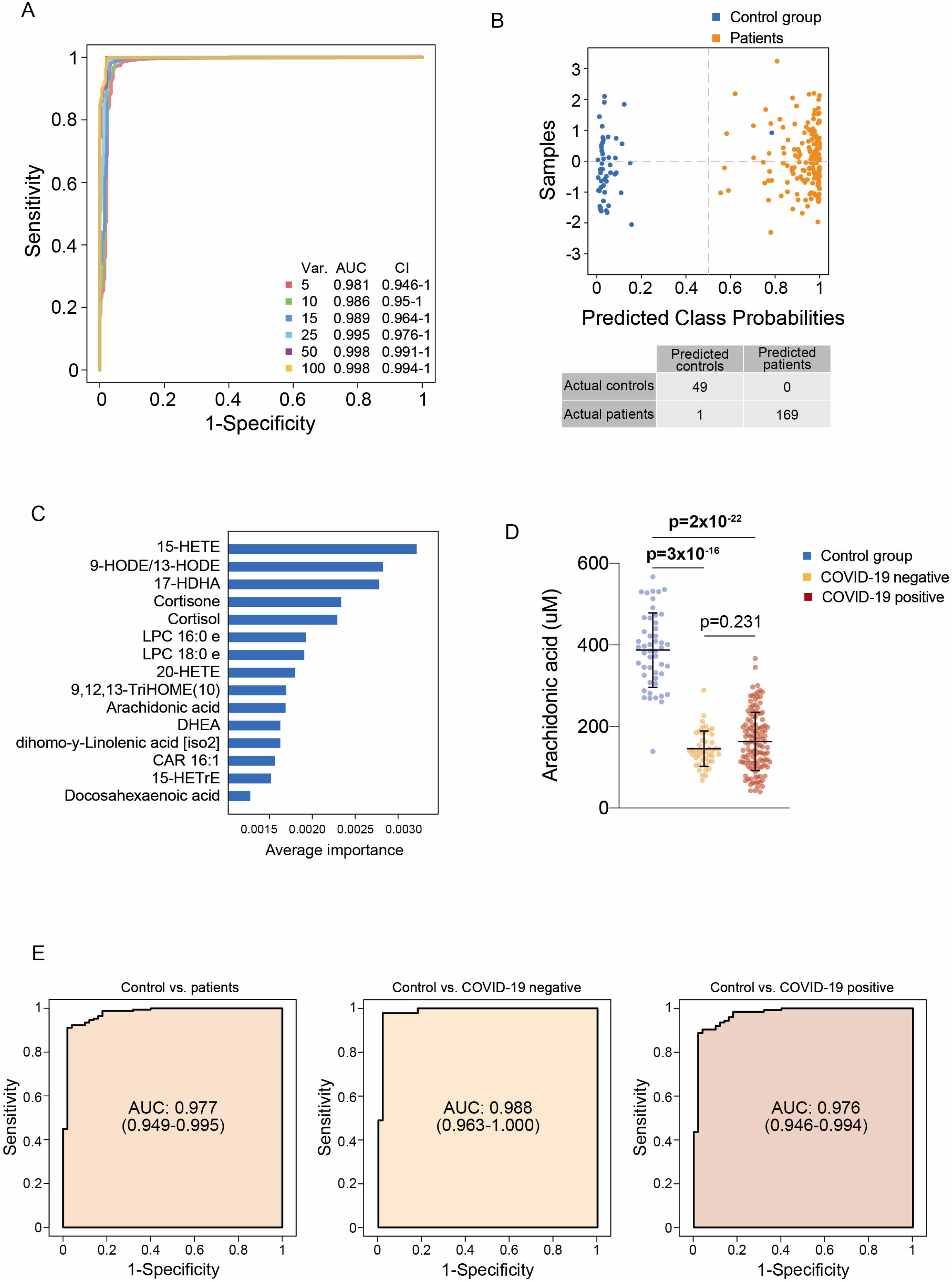
Identification of biomarkers for infectious/inflammatory processes. (A): Receiver Operating Characteristics plot of Monte Carlo models corresponding to the combination of 5 to 100 variables. (B): Confusion matrix of the generated 100-variable model. (C): Relative importance of the different variables chosen by the model. (D) Serum arachidonic acid concentrations in the COVID-19-positive and COVID-19-negative patients and the control group. (D): Receiver Operating Characteristics plots of the measured arachidonic acid in discriminating between the selected groups. Acronyms: AUC: Area under the curve; CAR: acylcarnitine; DHEA: dehydroepiandrosterone; HDHA: hydroxydocosahaxaenoic aid; HETE: hydroxyeicosatetraenoic acid; HODE: hydroxyoctadecadienoic acid; LPC: lysophosphatidylcholine; THOME: trihydroxyoctadecenoic acid.

### 3.3. Phosphatidylcholines and secondary bile acids are specifically altered in COVID-19 positive patients

Volcano plots identified changes in the concentrations of 86 species comparing the COVID-19-positive *vs*. COVID-19-negative patients (78 increased and 8 decreased in COVID-19-positive patients). The species that presented greatest changes were phosphatidylcholine 36:5 (PC 36:5, long-chain triglycerides (TG) 54:2 and 54:7 which were increased, and carnitine (CAR) 18:2, epoxystearic acid, and glycodeoxycholic acid, that were decreased in COVID-19-positive patients (Fig. 4A). PCA and PLS-DA showed separation but with a certain degree of overlap (Fig. 4B). The most relevant parameters in the discrimination between both groups of patients were the secondary bile acids deoxycholic acid and ursodeoxycholic/hyodeoxycholic acid (Fig. 4C). The heatmap clustered TG and PC values into two different groups, although with very similar behavior: they tended to be relatively more concentrated in COVID-19-positive patients than in the COVID-19-negative ones (Fig. 4D).

**Fig. 4.**
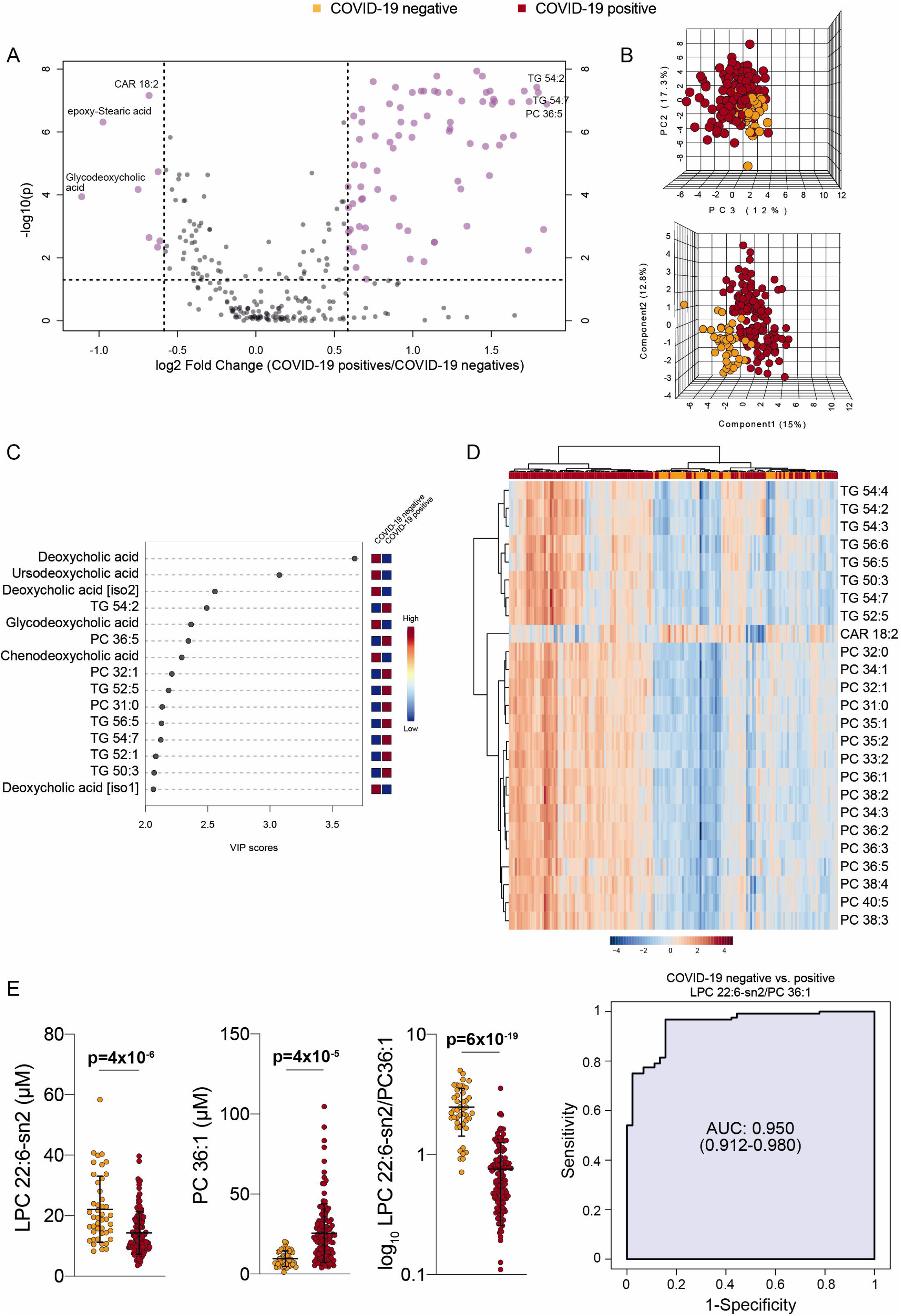
Lipid signatures differentiate between COVID-19-positive and COVID-19-negative patients. (A): Volcano plot representing the log fold-change of lipid species in COVID-19-positive with respect to COVID-19-negative patients. (B): Principal Component Analysis (PCA) clustering the COVID-19-positive and the COVID-19-negative patients. (C): Principal Least Square Discriminant Analysis (PLS-DA) clustering the COVID-19-positive and the COVID-19-negative patients. The Variable Importance in Projection (VIP) score identified deoxycholic and ursodeoxycholic/hyodeoxycholic acids as highly relevant parameters in the discrimination between both groups of patients. (D): Heatmap. (E): Serum concentrations of the selected lipid species in COVID-19-positive and COVID-19-negative patients, and Receiver Operating Characteristics plot of the ratio between them. In 3-dimensional plots of PCA and PLS-DA, each ball represents a patient, and position depends on differences in lipid concentrations. Axes are formed by different combination of variables, and percentages represent the proportion of variance that can be explained. PCA is a non-supervised test and PLS-DA is a supervised analysis. Acronyms: AUC: Area under the curve; CAR: acylcarnitine; LPC: lysophosphatidylcholine; PC: phosphatidylcholine; TG: triglyceride.

As in the previous section (described above), we generated Monte Carlo models to ascertain whether there was a biological marker that effectively discriminated between COVID-19-positive and COVID-19-negative patients (Supplementary Fig. 2A and B). The approach identified a variety of compounds, the concentrations of which differed in positive and negative patients (Supplementary Fig. 2C), but the AUC of even the best ROC curve did not exceed 0.8 (Supplementary Fig. 3D and 3E). Because the discriminatory ability of the Monte Carlo approach was modest, we manually tested the individual discriminatory ability of each of the variables; an AUC of 0.95 was obtained with the combination of LPC22:6-sn2 and PC36:1 (Fig. 4E).

### 3.4. Lipid profile in COVID-19-positive patients was related to specific comorbidities but not to clinical prognosis or survival

When we analyzed the lipid profile in relation to individual comorbidities, we observed several important differences. For example, patients with cancer had significantly higher levels of most lipid series than those patients without cancer, while patients with chronic lung disease had, in general, lower lipid levels (Fig. 5A). However, this analysis should be viewed with caution since most of the patients had more than one comorbidity and, as such, we prefer not to speculate on the influence of their interactions. To evaluate whether alterations in the lipid profile could be used to predict disease severity or mortality we applied K-means clustering in order to group patients according to their similarities within the circulating lipidome (Fig. 5B). All the distributions were dispersed and overlapped to a considerable extent, indicating that there was no significant relationship between lipid profile and survival, admission to the ICU, or the Charlson and McCabe indices.

**Fig. 5.**
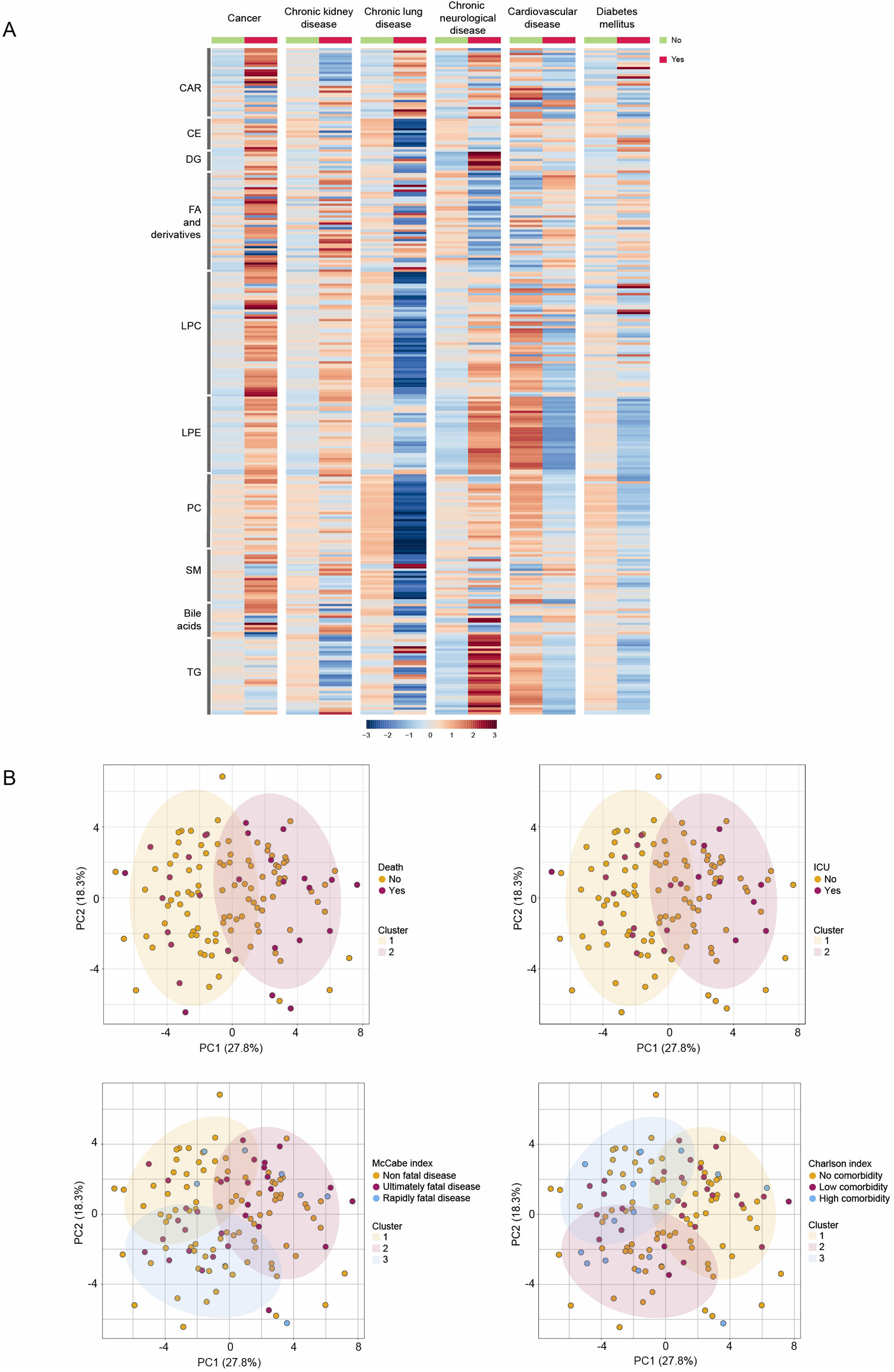
Relationships between the lipidomics signature and the clinical characteristics of COVID-19-positive patients. (A): Heatmap showing the variations in serum lipid concentrations in relation to comorbidities. (B): K-means clustering of patient group according to their similarities in the circulating lipidome. Each individual patient is represented by a point with a different color depending on whether or not they had the selected characteristic. Acronyms: CAR: acylcarnitines; CE: cholesterol esters; DG: diglycerides; FA: fatty acids; LPC: lysophosphatidylcholines; LPE: lysophosphatidylethanolamines; PC: phosphatidylcholines; SM: sphingomyelins; TG: triglycerides.

## 4. Discussion

When we compared the results of the COVID-19-positive patients with the healthy volunteers, the most relevant findings were the increases in the concentrations of CAR 8:0 and LPE, and the decrease in the concentrations of 9/13-HODE and 15-HETE. The enrichment analysis identified alterations in the synthesis pathway of arachidonic acid from fatty acids. The measurement of the serum levels of arachidonic acid showed a high level of discrimination between patients and control subjects. Increased serum CAR 8:0 concentrations in COVID-19-positive patients may be a reflection of mitochondrial dysfunction. Acylcarnitines are markers of mitochondrial function; specifically for β-oxidation of fatty acids. They are synthesized via carnitine palmitoyltransferase 1 that ferries fatty acids into the mitochondrial matrix. Incomplete fatty acid oxidation results in elevated acylcarnitine concentrations [20]. Indeed, our enrichment analysis suggested alterations in the pathways of mitochondrial β-oxidation of very-long-chain and medium-chain fatty acids. The mitochondrial long-chain fatty acids β-oxidation is impaired in several viral infections, including COVID-19 [21], while β-oxidation defects are mirrored by changes in the concentration of long-chain acylcarnitines. The accumulation of acylcarnitines within the lung has been reported to be a risk factor for acute lung injury due to their inhibition of pulmonary surfactants [22].

Fatty acids play essential roles in viral infection because they provide building blocks for membrane synthesis during virus proliferation, and also because fatty acids can be converted to many lipid mediators such as the eicosanoids, which play significant roles in immune and inflammatory responses [23]. We observed decreased serum concentrations of several fatty acids including arachidonic, stearic, lauric, and palmitic acid in COVID-19-positive patients compared with healthy individuals. This decrease may be related to enhanced synthesis pathways of viral membrane phospholipids. Among the fatty acids, the most marked alteration that we observed was a highly significant decrease in serum arachidonic acid concentration. This finding confirms an earlier study [24]. This may be relevant from a pathophysiological point of view in that arachidonic acid is a potent antiviral agent participating in the inactivation of enveloped viruses, including SARS-CoV-2 [10]. A decrease in the concentrations of this lipid would be detrimental to the host, and would encourage the survival of the invading virus. A further study reported that exogenous supplementation with arachidonic acid inhibited HcoV-229E virus replication in cultured cells [25]. The decrease in circulating levels of fatty acids was associated with a decrease in the concentrations of 9/13-HODE and 15-HETE; oxylipin products of the oxidation of linoleic acid and arachidonic acid, respectively. We would have expected to find increased serum oxylipin levels because their concentrations tend to increase by oxidative stress and because they are mediators of the inflammatory response [26]. However, an earlier study showed that high levels of oxylipins in lung cells infected by COVID-19 do not correspond to any concomitant increases in their concentrations in the circulation [27]. Indeed, these lipids are transported in plasma associated, mainly, with high-density lipoproteins from which they can be degraded by the antioxidant enzyme paraoxonase-1 [11,28].

Although the alterations in the lipid signature of COVID-19-positive patients are fairly unambiguous when compared to healthy subjects, COVID-19-negative patients presented similar alterations, as well. This finding suggests that these alterations were not specific to SARS-CoV-2 infection but, rather, are common to a multitude of infectious/inflammatory processes. For this reason, we compared the lipidomic signature of the COVID-19-positive patients with that of the COVID-19-negative ones. One alteration in particular was the significant difference in the circulating levels of phosphatidylcholines (PC) and lysophosphatidylcholines (LPC). Several studies have proposed a role of PC and LPC in COVID-19 infection, but the results published are far from consistent. Three studies had showed a decrease in plasma PC and an increase in LPC levels in COVID-19-positive patients compared to healthy subjects [29-31] while others showed that both phospholipids decreased [32,33], or even that the concentrations of PC increased [6] and those of LPC decreased [34]. We found a decrease in the serum concentration of LPC 22:6 and an increase in that of PC 36:1 and, hence, the ratio between the two phospholipids discriminated fairly well between positive and negative patients, and with excellent diagnostic accuracy. In addition, Volcano plots identified PC 36:5 as one of the lipid species that was most strongly increased when comparing positive *vs*. negative patients. These results agree with those reported in Calu-3 cells, where an increase in PC synthesis was observed when the cells were infected with SARS-CoV-2 [35]. Differences between the characteristics of the lipid groups studied can probably explain this discrepancy between different authors’ findings. Indeed, we need to highlight that, from amongst these studies, ours is unique in comparing COVID-19 patients with patients with infectious/inflammatory diseases of origins other than COVID-19 infection. Several factors could influence plasma PC and LPC concentrations. For example, both are key components of cell membranes and lipoproteins, and low plasma levels of these compounds may be explained as resulting from liver impairment in patients with severe COVID-19, while their increase would suggest increased activity of phospholipase A2 [35]. Alterations in PC and LPC levels have been related to disease severity because of the role that these lipids play in the inflammatory response [36].

Another alteration that we observed in the COVID-19-positive patients when compared with the COVID-19-negative ones was a decrease in the concentrations of secondary bile acids, mainly deoxycholic acid and ursodeoxycholic/hyodeoxycholic acid; products of metabolism in the human gut microbiome. Our results are in concordance with those reporting that the fecal microbiome diversity is decreased in COVID-19 patients [37] and SARS-CoV-2-infected primates [9]. Moreover, decreased plasma deoxycholic concentrations have been reported in severe COVID-19 patients compared to those with milder forms of the disease [38]. A disruption in the interaction between the gut and the lung has been related to respiratory tract diseases, including the acute respiratory distress syndrome [39]. Inflammation caused by lung infection can disrupt the gut barrier integrity and increase the permeability to gut microbes and microbial products. This microbial translocation can exacerbate inflammation resulting from positive feedback. Further, microbial translocation may also modulate the circulating levels of gut microbiota-associated products such as secondary bile acids. As such, the circulating levels of these compounds would reflect the functional status of the gut and the metabolic activity of its microbiota [40]. Also, they are biologically active molecules that regulate several immunological functions, including inflammatory responses. Indeed, ursodeoxycholic acid has antioxidant, anti-inflammatory, anti-apoptotic, and immunomodulatory properties [8].

We did not find any significant difference in the lipidomic signature of patients who survived and those who did not, nor with admission to the ICU, nor in the clinical prognosis. In this sense we differ from earlier studies, albeit the published information is scarce. For example, Siendelar et al. [36] found that a panel of 22 metabolites (including PC and LPC), predicted disease severity (as measured as need for ICU admission). Giron et al. [38] reported that alterations in secondary bile acids levels, resulting from disrupted crosstalk between gut and lung, are associated with ICU admission. The reasons for these discrepancies are purely speculative. Plausible explanations could be the heterogeneity of the disease itself, the different levels of severity and, as well, the associated comorbidities in the groups of patients studied by the different authors.

In summary, we identified CAR, LPE, arachidonic acid and oxylipins as the most altered parameters in COVID-19 patients compared to healthy volunteers. However, our study is also a cautionary note in that it shows these alterations to be not specific to COVID-19, and occur in other diseases with an infectious/inflammatory component. We also identified long-chain TG, LPC22:6-sn2, PC36:1 and secondary bile acids as the parameters that present the greatest capacity to discriminate between positive and negative COVID-19 hospitalized patients. These lipid alterations highlight the option of continuing to treat these patients post-discharge from hospital. Given the pro-atherogenic role of some of these lipid species, follow-up treatment could include lifestyle modifications and lipid-lowering drugs. Our systematic investigation showed that the integration of lipidomics with machine learning algorithms can increase the understanding of COVID-19 pathophysiology and, as such, facilitate more effective clinical decision making.

## Data Availability

All data produced in the present study are available upon reasonable request to the authors

## CRediT authorship contribution statement

**Helena Castañé**: Conceptualization, Methodology, Software, Formal analysis, Investigation, Data curation, Writing-Original draft preparation. **Simona Iftimie:** Conceptualization, Methodology, Formal analysis, Investigation, Data curation, Writing-Original draft preparation. **Gerard Baiges-Gaya**: Methodology, Investigation. **Elisabet Rodríguez-Tomàs:** Methodology, Investigation. **Andrea Jiménez-Franco**: Methodology, Investigation. **Ana Felisa López-Azcona:** Formal analysis, Investigation, Data curation. **Pedro Garrido:** Formal analysis, Investigation, Data curation. Antoni **Castro:** Data curation, Funding acquisition. **Jordi Camps**: Conceptualization, Formal analysis, Supervision, Project administration, Writing-Review & Editing. **Jorge Joven:** Data curation, Funding acquisition.

## Declaration of competing interest

The authors report no potential conflicts of interest relevant to this article.

## Acknowledgments

The authors thank Dr. Peter R. Turner for critical review and language editing of the manuscript. The EURECAT-Technology Centre of Catalonia (Reus, Spain) contributed its expertise and advice in lipidomics measurements.

## Funding

This work was supported by the *Fundació La Marató de TV3*, Barcelona, Spain [201807-10], the *Generalitat de Catalunya*, Barcelona, Spain [PERIS-SLT017/3501/2020 to H.C. and AGAUR 2020FI_d1 00215 to E.R.T.], and the *Instituto de Salud Carlos* III, Madrid, Spain [FI19/00097 to G.B.G.]. Funders had no role in the study design, data collection, data analysis, data interpretation, and writing the manuscript.

## Notes

### Competing Interest Statement

The authors have declared no competing interest.

### Funding Statement

This work was supported by the Fundacio La Marato de TV3, Barcelona, Spain [201807-10], the Generalitat de Catalunya, Barcelona, Spain [PERIS-SLT017/3501/2020 to H.C. and AGAUR 2020FI_d1 00215 to E.R.T.], and the Instituto de Salud Carlos III, Madrid, Spain [FI19/00097 to G.B.G.]. Funders had no role in the study design, data collection, data analysis, data interpretation, and writing the manuscript.

### Author Declarations

This study was approved by the Comite de Etica i Investigacio en Medicaments (Institutional Review Committee) of the Institut deInvestigacio Sanitaria Pere Virgili (Resolution CEIM 040/2018, modified on April 16, 2020).

