## Supplementary Material for "Machine learning identified distinct serum lipidomic signatures in hospitalized COVID-19-positive and COVID-19-negative patients"

**Short title:** Serum lipidomics in COVID-19

**Keywords:** artificial intelligence; COVID-19; lipid metabolism; lipidomics; machine learning

**INDEX:**

**Page**

Supplementary Methods 1 – Lipidomics analysis	3

Supplementary Methods 2 – Machine learning analysis	4

Supplementary Figure 1	13

Supplementary Figure 2	14

Supplementary Methods 1 – Lipidomics analysis:

Three different analytical methods were used to quantify 283 lipid species:

Acylcarnitines:

L-carnitine, O-acetyl-L-carnitine, O-propionyl-L-carnitine, O-butyryl-L-carnitine, O-isovaleryl-L-carnitine, O-myristoyl-L-carnitine, and O-palmitoyl-L-carnitine (Cambridge Isotope Laboratories Inc., Tewksbury, MA, USA), and O-glutaryl-L-carnitine, O-3-hydroxyisovaleryl-L-carnitine, O-dodeanoyl-L-carnitine, and O-3-DL-hydroxypalmitoyl-L-carnitine (Cambrige Isotope Laboratories Inc.) were used as analytical standards. L-carnitine, O-acetyl-L-carnitine, O-propionyl-L-carnitine, O-butyryl-L-carnitine, O-isovaleryl-L-carnitine, O-myristoyl-L-carnitine, and O-palmitoyl-L-carnitine (Cambridge Isotope Laboratories Inc.) were used as internal standards.

Fifty μL of serum were thawed at 4ºC and mixed with 200 μL of methanol containing the set of labeled internal standards. The mixture was vortexed for 15 seconds and centrifuged for 10 min at 20,000xg at 4ºC. The supernatant was transferred into a chromatographic vial and injected on liquid chromatography coupled to mass spectrometry (LC-MS/MS) (UHPLC 1290 Infinity II Series coupled to a QqQ/MS 6470 Series, Agilent Technologies, Santa Clara, CA, USA).

The mobile phase consisted of A: 99.9% water + 0.1% formic acid; B: 99.9% methanol with 0.1% formic acid at a flow rate of 0.4 mL/min. The gradient used was as follows: 0 min, 0% B; 11 min, 100% B, 13 min, 0%B, 16.5 min, 0%B. The column (Kinetex 2.6 μm Polar C18, 100 Å, 150 x 2.1 mm, Phenomenex, Torrance, CA, USA) temperature was set at 20ºC and the injection volume was 1 μL. Electrospray ionization source and triple quadrupole mass parameters are: Ionization mode, positive; gas temperature, 200ºC; gas flow, 11 L/min; nebulizer, 30 psi; sheath gas temperature, 375ºC; sheath gas flow, 11 L/min; capillary voltage, 3,500 V.

Non-polar lipids:

Lysophosphatidylcholine (LPC) 18:0, phosphatidylcholine (PC) 32:0, sphingomyelin (SM) 36:1, diglyceride (DG) 36:0, triglyceride (TG) 52:3, and cholesterol ester (CE) 16:0 were used as analytical standards (Avanti Polar Lipids, Alabaster, AL, USA), and LPC 18:1-d7, PC 33:1-d7, SM 36:2-d9, DG 33:1-d7, TG 48:1-d7, and CE 18:1-d7 were used as internal standards (SPLASH, Avanti Polar Lipids).

Twenty μL of serum were mixed with 20 μL of NaCl 0.8% and vortexed. Then, 200 μL of CHCl_3_:MeOH 2:1 were added, samples were vortexed again and centrifuged at 20,000xg at 4ºC for 10 min. The lower phase was recovered, evaporated to dryness and reconstituted with methanol:methyl-tert-butyl ether (9:1) and analyzed by UHPLC-ESI-qTOF-MS (UHPLC 1290 Infinity II Series coupled to a qTOF/MS 6550 Series, Agilent Technologies).

The chromatographic method consisted in an elution with a ternary mobile phase containing water (A), methanol (B) and 2-propanol with 10mM ammonium formate and formic acid 0.1% (C) at a flow rate of 0.6 mL/min and the following gradient program: 0 min, 10% B, 45% C; 0.5 min, 9.5% B, 45.0% C; 1.5 min, 7.5% B, 47.7% C; 1.6 min, 7% B, 58.5% C; 5 min, 4% B, 61.2% C; 5.1 min, 3.5% B, 77.4% C; 7.5 min, 3.5% B, 80.1% C; 9 min, 0% B, 80.1% C; 9 min, 0% B, 80.1% C; 9.5 min, 0% B, 100% C; 11.5 min, 10% B, 100% C). The stationary phase, thermostatized at 60ºC, was a Kinetex EVO C18 column (2.6 μm, 2.1 mm x 100 mm). Optimized parameters for ESI-qTOF-MS are: Ionization mode, positive; gas temperature, 225ºC; gas flow, 11 L/min; nebulizer, 35 psi; sheath gas temperature, 300ºC; sheath gas flow, 12 L/min; capillary voltage, 3,500 V, nozzle voltage, 500 V.

**Polar lipids:**

Lysophosphatidylethanolamine (LPE) 16:0, LPC 18:0, Dehydroepiandrosterone 3-sulfate, cortisol, cholic acid, taurocholic acid, deoxycholic acid, arachidonic acid, and 15-hydroxyeicosatetraenoic acid (15-HETE) were used as standards, and LPC 18:1-d7, cholic acid-d4, taurocholic acid-d5, arachidonic acid-d8, and myristic acid-d27 were used as internal standards.

Fifty μL of serum were thawed at 4ºC and mixed with 200 μL of methanol containing the set of labeled internal standards. Then, samples were vortexed and centrifuged at 20,000xg for 10 minutes at 4ºC. The supernatant was analyzed by UHPLC-qTOF (UHPLC 1290 Infinity II Series coupled to a qTOF/MS 6550 Series, both Agilent Technologies).

The chromatographic method consisted in an elution with water (A) and acetonitrile (B), both with formic acid 0.05%, as a mobile phase. The gradient used was: 0 min, 0% B; 11 min, 100% B, 13 min, 0%B, 16.5 min, 0%B. As stationary phase, an ACQUITY BEH C18 Column (1.7 μm, 2.1 mm x 100 mm) was employed. ESI-q-TOF-MS parameters are: Ionization mode, positive; gas temperature, 225ºC; gas flow, 11 L/min; nebulizer, 35 psi; sheath gas temperature, 300ºC; sheath gas flow, 12 L/min; capillary voltage, 3,500 V, nozzle voltage, 500 V.

Lipid identification:

Lipid identification was performed by matching their accurate mass and tandem mass spectrum to Metlin-PCDL from Agilent when available. In addition, the chromatographic behavior of pure standards for each family and bibliographic information were used to ensure their putative identification. The identification indicates the lipid family, the total number of carbons of the acyl chains, and the number of double bonds. Internal standards were used to correct the response of each detected lipid species. The obtained calibration curves were used for the quantification of their corresponding lipid species. For the rest of compounds, the analysis was semi-quantitative using the appropriate species for each family of lipid.

**Supplementary Methods 2 – Machine learning analysis:**

Below, we show the R-command histories of the MetaboAnalystR for the different types of analysis.

Groups are identified as: 1, control group; 2, COVID-19 negative; 3, COVID-19 positive.

**Univariate statistics:**

library(tableone)
catVars <- c("sex", "smoking", "alcoh", "c_icu", "c_death", "s_fever", "s_cough", "s_pneum", "s_odynop", "s_chills", "s_resdis", "s_vom", "s_ards", "s_diarrh", "s_akf", "s_other", "co_dm", "co_cvd", "co_cld", "co_clud", "co_ckd", "co_cnd", "co_inf", "co_canc", "co_preg", "co_postp", "r_nimv", "r_imv", "r_hfot", "r_cot", "r_ri", "co_charl", "co_mccab", "cs_il10q", "cs_il6q", "d_insul", "d_stat")
myVars <- c("sex", "age", "smoking", "alcoh", "cs_ccl2",
"cs_gal3", "cs_pon1a", "cs_pon1c", "cs_il1", "cs_il6", "cs_ag2",
"cs_ace2", "bt_crp", "bt_gluc", "bt_creat", "bt_chol", "bt_hdlc",
"bt_ldlc", "bt_vldlc", "bt_tg", "bt_ast", "bt_alt", "bt_hb",
"bt_eryt", "bt_leuk", "bt_neutr", "bt_lymph", "bt_monoc", "bt_eosin",
"bt_basop", "bt_plat", "c_icu", "c_days", "c_death", "s_fever",
"s_cough", "s_pneum", "s_odynop", "s_chills", "s_resdis", "s_vom",
"s_ards", "s_diarrh", "s_akf", "s_other", "co_dm", "co_cvd",
"co_cld", "co_clud", "co_ckd", "co_cnd", "co_inf", "co_canc",
"co_preg", "co_postp", "r_nimv", "r_imv", "r_hfot", "r_cot",
"r_ri", "co_charl", "co_mccab", "cs_il10q", "cs_il6q", "d_insul",
"d_stat", "CAR0", "CAR10_0", "CAR10_1", "CAR12_0", "CAR12_1",
"CAR13_0", "CAR14_0", "CAR14_1", "CAR14_2", "CAR15_0", "CAR16_0",
"CAR16_1", "CAR16_2", "CAR18_0", "CAR18_1", "CAR18_2", "CAR3_0",
"CAR3_1", "CAR4_0", "CAR4_0.2M", "CAR5_0", "CAR5_1", "CAR5.DC",
"CAR5.M.DC", "CAR6_0", "CAR7_0", "CAR8_0", "CAR8_0.DC", "CAR8_1",
"CAR9_0", "CE16_0", "CE16_1", "CE17_1", "CE18_0", "CE18_1", "CE18_2",
"CE18_3", "CE20_2", "CE20_3", "CE20_4", "CE20_5", "CE22_4", "CE22_5",
"CE22_6", "DG34_1", "DG34_2", "DG34_3", "DG36_0", "DG36_1", "DG36_2",
"DG36_3", "DG36_4", "DG40_4", "FA12_0", "FA14_0", "FA16_0", "FA16_1c",
"FA16_1t", "FA18_0", "FA18_1", "FA18_1O", "FA18_1O2.1", "FA18_1O2.2",
"FA18_1O2h", "FA18_1O3", "FA18_2", "FA18_2O", "FA18_2O2", "FA18_2OEp",
"FA18_2OEp9", "FA18_3i1", "FA18_3i2", "FA18_3O2", "FA18_3Oi1",
"FA18_4", "FA18_4i1", "FA19_1", "FA20_0", "FA20_2", "FA20_3i1",
"FA20_3i2", "FA20_3i3", "FA20_3O", "FA20_4", "FA20_4O12", "FA20_4O15",
"FA20_4O20", "FA20_4Ox", "FA20_5", "FA20_5O2", "FA22_4", "FA22_5w3",
"FA22_5w6", "FA22_6", "FA22_6O", "LPC14_0sn1", "LPC14_0sn2",
"LPC15_0", "LPC15_0sn1", "LPC15_0sn2", "LPC16_0", "LPC16_0e",
"LPC16_0sn1", "LPC16_0sn2", "LPC16_1e", "LPC16_1sn1", "LPC16_1sn2",
"LPC17_0sn1", "LPC17_0sn2", "LPC17_1sn1", "LPC17_1sn2", "LPC18_0",
"LPC18_0e", "LPC18_0sn1", "LPC18_0sn2", "LPC18_1", "LPC18_1sn1",
"LPC18_1sn2", "LPC18_2", "LPC18_2sn1", "LPC18_2sn2", "LPC18_3sn1",
"LPC18_3sn2", "LPC19_0sn1", "LPC19_0sn2", "LPC20_0", "LPC20_0sn1",
"LPC20_0sn2", "LPC20_1sn1", "LPC20_1sn2", "LPC20_2", "LPC20_2sn1",
"LPC20_2sn2", "LPC20_3", "LPC20_3sn1", "LPC20_3sn2", "LPC20_4sn1",
"LPC20_4sn2", "LPC20_5sn1", "LPC20_5sn2", "LPC22_3sn1", "LPC22_3sn2",
"LPC22_4sn1", "LPC22_4sn2", "LPC22_5sn1", "LPC22_5sn2", "LPC22_6sn1",
"LPC22_6sn2", "LPE14_0sn1", "LPE14_0sn2", "LPE15_0sn1", "LPE15_0sn2",
"LPE16_0sn1", "LPE16_0sn2", "LPE16_1sn1", "LPE16_1sn2", "LPE17_0sn1",
"LPE17_1sn1", "LPE17_1sn2", "LPE18_0sn1", "LPE18_0sn2", "LPE18_1sn1",
"LPE18_1sn2", "LPE18_2sn1", "LPE18_2sn2", "LPE18_3sn1", "LPE18_3sn2",
"LPE20_1sn1", "LPE20_1sn2", "LPE20_2sn1", "LPE20_3sn1", "LPE20_3sn2",
"LPE20_4sn1", "LPE20_4sn2", "LPE20_5sn1", "LPE20_5sn2", "LPE22_4sn1",
"LPE22_4sn2", "LPE22_5sn1", "LPE22_6sn1", "LPE22_6sn2", "NAE16_0",
"NAE18_0", "NAE18_1", "NAE18_2", "PC30_0", "PC31_0", "PC32_0",
"PC32_1", "PC32_2", "PC33_0", "PC33_1", "PC33_2", "PC34_0", "PC34_1",
"PC34_2", "PC34_3", "PC34_4", "PC35_1", "PC35_2", "PC35_4", "PC36_1",
"PC36_2", "PC36_3", "PC36_4", "PC36_5", "PC38_2", "PC38_3", "PC38_4",
"PC38_5", "PC38_6", "PC40_4", "PC40_5", "SM32_1", "SM32_2", "SM33_1",
"SM34_1", "SM34_2", "SM35_1", "SM36_0", "SM36_1", "SM36_2", "SM38_1",
"SM38_2", "SM39_1", "SM40_1", "SM40_2", "SM41_1", "SM41_2", "SM42_1",
"SM42_2", "SM42_3", "SM43_1", "SM43_2", "SPBP16_0O2", "SPBP18_1O2",
"ST19_2O2", "ST21_3O5", "ST21_4O5", "ST24_1O3T", "ST24_1O4",
"ST24_1O4C", "ST24_1O4G", "ST24_1O4Gc", "ST24_1O4Gg", "ST24_1O4i1",
"ST24_1O4i2", "ST24_1O4u", "ST24_1O5", "ST24_1O5G", "ST24_1O5T",
"TG46_0", "TG46_1", "TG46_2", "TG47_0", "TG47_1", "TG48_0", "TG48_1",
"TG48_2", "TG48_3", "TG50_0", "TG50_1", "TG50_2", "TG50_3", "TG50_4",
"TG51_2", "TG51_3", "TG52_1", "TG52_2", "TG52_3", "TG52_4", "TG52_5",
"TG52_6", "TG54_2", "TG54_3", "TG54_4", "TG54_5", "TG54_6", "TG54_7",
"TG56_5", "TG56_6", "TG56_7", "TG58_8")
tab1 <- CreateTableOne(vars = myVars, strata = "group" , data = COVID_Lipidomics_and_clinics, factorVars = catVars)
print(tab1, showAllLevels = TRUE, formatOptions = list(big.mark = ","), quote = TRUE, noSpaces = TRUE)

**Multivariate statistics:**

From the R package MetaboAnalystR we generated csv files to list P-values and FDR. We also generated the following plots for each comparison: Volcano plot, PCA loading plot, PLS-DA loading plot, PLS-DA VIP scores, PLS-DA cross validation, and clustered and not-clustered heatmaps.

library(MetaboAnalystR)

#Ctrl vs. COVID-19-negative vs. COVID-19-positive
mSet<-InitDataObjects("conc", "stat", FALSE)
mSet<-Read.TextData(mSet, "COVID_Lipidomics.csv", "rowu", "disc");
mSet<-SanityCheckData(mSet)
mSet<-ContainMissing(mSet)
mSet<-SanityCheckData(mSet)
mSet<-ContainMissing(mSet)
mSet<-RemoveMissingPercent(mSet, percent=0.5)
mSet<-ImputeMissingVar(mSet, method="knn_var")
mSet<-SanityCheckData(mSet)
mSet<-ContainMissing(mSet)
mSet<-FilterVariable(mSet, "none", "F", 25)
mSet<-PreparePrenormData(mSet)
mSet<-Normalization(mSet, "NULL", "LogNorm", "NULL", ratio=FALSE, ratioNum=20)
mSet<-PCA.Anal(mSet)
mSet<-PlotPCAPairSummary(mSet, "pca_pair_0_", "pdf", 72, width=NA, 5)
mSet<-PlotPCAScree(mSet, "pca_scree_0_", "pdf", 72, width=NA, 5)
mSet<-PlotPCA2DScore(mSet, "pca_score2d_0_", "pdf", 72, width=NA, 1,2,0.95,0,0)
mSet<-PlotPCALoading(mSet, "pca_loading_0_", "pdf", 72, width=NA, 1,2);
mSet<-PlotPCABiplot(mSet, "pca_biplot_0_", "pdf", 72, width=NA, 1,2)
mSet<-PlotPCA3DLoading(mSet, "pca_loading3d_0_", "json", 1,2,3)
mSet<-GetGroupNames(mSet, "null")
mSet<-PlotPCA2DScore(mSet, "pca_score2d_0_", "pdf", 72, width=NA, 1,2,0.95,0,0)
mSet<-SaveTransformedData(mSet)
mSet<-PlotPCAPairSummary(mSet, "pca_pair_0_", "pdf", 72, width=NA, 5)
mSet<-PLSR.Anal(mSet, reg=TRUE)
mSet<-PlotPLSPairSummary(mSet, "pls_pair_0_", "pdf", 72, width=NA, 5)
mSet<-PlotPLS2DScore(mSet, "pls_score2d_0_", "pdf", 72, width=NA, 1,2,0.95,0,0)
mSet<-PlotPLS3DScoreImg(mSet, "pls_score3d_0_", "pdf", 72, width=NA, 1,2,3, 40)
mSet<-PlotPLSLoading(mSet, "pls_loading_0_", "pdf", 72, width=NA, 1, 2);
mSet<-PlotPLS3DLoading(mSet, "pls_loading3d_0_", "json", 1,2,3)
mSet<-PLSDA.CV(mSet, "T",5, "Q2")
mSet<-PlotPLS.Classification(mSet, "pls_cv_0_", "pdf", 72, width=NA)
mSet<-PlotPLS.Imp(mSet, "pls_imp_0_", "pdf", 72, width=NA, "vip", "Comp. 1", 15,FALSE)
mSet<-PlotPLSPairSummary(mSet, "pls_pair_0_", "pdf", 72, width=NA, 5)
mSet<-PlotPLS2DScore(mSet, "pls_score2d_0_", "pdf", 72, width=NA, 1,2,0.95,0,0)
mSet<-PlotPLS.Imp(mSet, "pls_imp_0_", "pdf", 72, width=NA, "vip", "Comp. 1", 15,FALSE)
mSet<-PlotPLS.Classification(mSet, "pls_cv_0_", "pdf", 72, width=NA)
mSet<-PlotHeatMap(mSet, "heatmap_0_", "pdf", 72, width=NA, "norm", "row", "euclidean", "ward.D","bwm", "overview", T, T, NULL, T, F)
mSet<-PlotHeatMap(mSet, "heatmap_1_", "pdf", 72, width=NA, "norm", "row", "euclidean", "ward.D","bwm", "detail", T, T, NULL, F, F)
mSet<-PlotHeatMap(mSet, "heatmap_3_", "pdf", 72, width=NA, "norm", "row", "euclidean", "ward.D","bwm", "detail", F, F, NULL, F, F)
mSet<-PlotSubHeatMap(mSet, "heatmap_4_", "pdf", 72, width=NA, "norm", "row", "euclidean", "ward.D","bwm", "tanova", 25, "overview", T, T, F, F)
mSet<-PlotSubHeatMap(mSet, "heatmap_5_", "pdf", 72, width=NA, "norm", "row", "euclidean", "ward.D","bwm", "vip", 15, "overview", T, T, F, T)

#COVID-19-negative vs. COVID-19-positive
feature.nm.vec <- c("")
smpl.nm.vec <- c("")
grp.nm.vec <- c("1")
mSet<-UpdateData(mSet)
mSet<-PreparePrenormData(mSet)
mSet<-Normalization(mSet, "NULL", "LogNorm", "NULL", ratio=FALSE, ratioNum=20)
mSet<-PlotNormSummary(mSet, "norm_1_", "png", 72, width=NA)
mSet<-PlotSampleNormSummary(mSet, "snorm_1_", "png", 72, width=NA)
mSet<-FC.Anal(mSet, 2.0, 0, FALSE)
mSet<-PlotFC(mSet, "fc_0_", "png", 72, width=NA)
mSet<-FC.Anal(mSet, 2.0, 1, FALSE)
mSet<-PlotFC(mSet, "fc_1_", "png", 72, width=NA)
mSet<-FC.Anal(mSet, 1.0, 1, FALSE)
mSet<-PlotFC(mSet, "fc_2_", "png", 72, width=NA)
mSet<-Ttests.Anal(mSet, F, 0.05, FALSE, TRUE, "fdr", FALSE)
mSet<-PlotTT(mSet, "tt_0_", "png", 72, width=NA)
mSet<-Ttests.Anal(mSet, T, 1.0, FALSE, TRUE, "fdr", FALSE)
mSet<-PlotTT(mSet, "tt_1_", "png", 72, width=NA)
mSet<-Volcano.Anal(mSet, FALSE, 2.0, 0, F, 0.1, TRUE, "raw")
mSet<-PlotVolcano(mSet, "volcano_0_",1, "png", 72, width=NA)
mSet<-Volcano.Anal(mSet, FALSE, 1.5, 1, T, 0.05, TRUE, "fdr")
mSet<-PlotVolcano(mSet, "volcano_1_",1, "png", 72, width=NA)
mSet<-PlotVolcano(mSet, "volcano_1_",1, "pdf", 72, width=NA)
mSet<-PCA.Anal(mSet)
mSet<-PlotPCAPairSummary(mSet, "pca_pair_0_", "png", 72, width=NA, 5)
mSet<-PlotPCAScree(mSet, "pca_scree_0_", "png", 72, width=NA, 5)
mSet<-PlotPCA2DScore(mSet, "pca_score2d_0_", "png", 72, width=NA, 1,2,0.95,0,0)
mSet<-PlotPCALoading(mSet, "pca_loading_0_", "png", 72, width=NA, 1,2);
mSet<-PlotPCABiplot(mSet, "pca_biplot_0_", "png", 72, width=NA, 1,2)
mSet<-PlotPCA3DLoading(mSet, "pca_loading3d_0_", "json", 1,2,3)
mSet<-GetGroupNames(mSet, "null")
mSet<-PlotPCAPairSummary(mSet, "pca_pair_0_", "pdf", 72, width=NA, 5)
mSet<-PlotPCA2DScore(mSet, "pca_score2d_0_", "pdf", 72, width=NA, 1,2,0.95,0,0)
mSet<-PLSR.Anal(mSet, reg=TRUE)
mSet<-PlotPLSPairSummary(mSet, "pls_pair_0_", "png", 72, width=NA, 5)
mSet<-PlotPLS2DScore(mSet, "pls_score2d_0_", "png", 72, width=NA, 1,2,0.95,0,0)
mSet<-PlotPLS3DScoreImg(mSet, "pls_score3d_0_", "png", 72, width=NA, 1,2,3, 40)
mSet<-PlotPLSLoading(mSet, "pls_loading_0_", "png", 72, width=NA, 1, 2);
mSet<-PlotPLS3DLoading(mSet, "pls_loading3d_0_", "json", 1,2,3)
mSet<-PLSDA.CV(mSet, "T",5, "Q2")
mSet<-PlotPLS.Classification(mSet, "pls_cv_0_", "png", 72, width=NA)
mSet<-PlotPLS.Imp(mSet, "pls_imp_0_", "png", 72, width=NA, "vip", "Comp. 1", 15,FALSE)
mSet<-PlotPLSPairSummary(mSet, "pls_pair_0_", "pdf", 72, width=NA, 5)
mSet<-PlotPLS2DScore(mSet, "pls_score2d_0_", "pdf", 72, width=NA, 1,2,0.95,0,0)
mSet<-PlotPLS.Imp(mSet, "pls_imp_0_", "pdf", 72, width=NA, "vip", "Comp. 1", 15,FALSE)
mSet<-PlotPLS.Classification(mSet, "pls_cv_0_", "pdf", 72, width=NA)
mSet<-PlotHeatMap(mSet, "heatmap_5_", "png", 72, width=NA, "norm", "row", "euclidean", "ward.D","bwm", "overview", T, T, NULL, T, F)
mSet<-PlotHeatMap(mSet, "heatmap_6_", "png", 72, width=NA, "norm", "row", "euclidean", "ward.D","bwm", "detail", T, T, NULL, F, F)
mSet<-PlotHeatMap(mSet, "heatmap_6_", "pdf", 72, width=NA, "norm", "row", "euclidean", "ward.D","bwm", "detail", T, T, NULL, F, F)
mSet<-PlotHeatMap(mSet, "heatmap_7_", "png", 72, width=NA, "norm", "row", "euclidean", "ward.D","bwm", "detail", F, F, NULL, F, F)
mSet<-PlotHeatMap(mSet, "heatmap_7_", "pdf", 72, width=NA, "norm", "row", "euclidean", "ward.D","bwm", "detail", F, F, NULL, F, F)
mSet<-PlotSubHeatMap(mSet, "heatmap_8_", "png", 72, width=NA, "norm", "row", "euclidean", "ward.D","bwm", "tanova", 25, "overview", T, T, F, F)
mSet<-PlotSubHeatMap(mSet, "heatmap_8_", "pdf", 72, width=NA, "norm", "row", "euclidean", "ward.D","bwm", "tanova", 25, "overview", T, T, F, F)
mSet<-PlotSubHeatMap(mSet, "heatmap_9_", "png", 72, width=NA, "norm", "row", "euclidean", "ward.D","bwm", "tanova", 15, "overview", T, T, F, T)
mSet<-PlotSubHeatMap(mSet, "heatmap_9_", "pdf", 72, width=NA, "norm", "row", "euclidean", "ward.D","bwm", "tanova", 15, "overview", T, T, F, T)
mSet<-PlotSubHeatMap(mSet, "heatmap_10_", "png", 72, width=NA, "norm", "row", "euclidean", "ward.D","bwm", "vip", 15, "overview", T, T, F, T)
mSet<-PlotSubHeatMap(mSet, "heatmap_10_", "pdf", 72, width=NA, "norm", "row", "euclidean", "ward.D","bwm", "vip", 15, "overview", T, T, F, T)

#Ctrl vs COVID-19 positives
feature.nm.vec <- c("")
smpl.nm.vec <- c("")
grp.nm.vec <- c("2")
mSet<-UpdateData(mSet)
mSet<-PreparePrenormData(mSet)
mSet<-Normalization(mSet, "NULL", "LogNorm", "NULL", ratio=FALSE, ratioNum=20)
mSet<-PlotNormSummary(mSet, "norm_2_", "png", 72, width=NA)
mSet<-PlotSampleNormSummary(mSet, "snorm_2_", "png", 72, width=NA)
mSet<-FC.Anal(mSet, 2.0, 0, FALSE)
mSet<-PlotFC(mSet, "fc_2_", "png", 72, width=NA)
mSet<-FC.Anal(mSet, 1.0, 1, FALSE)
mSet<-PlotFC(mSet, "fc_3_", "png", 72, width=NA)
mSet<-Ttests.Anal(mSet, F, 0.05, FALSE, TRUE, "fdr", FALSE)
mSet<-PlotTT(mSet, "tt_1_", "png", 72, width=NA)
mSet<-Ttests.Anal(mSet, T, 1.0, FALSE, TRUE, "fdr", FALSE)
mSet<-PlotTT(mSet, "tt_2_", "png", 72, width=NA)
mSet<-Volcano.Anal(mSet, FALSE, 2.0, 0, F, 0.1, TRUE, "raw")
mSet<-PlotVolcano(mSet, "volcano_1_",1, "png", 72, width=NA)
mSet<-Volcano.Anal(mSet, FALSE, 1.5, 1, T, 0.05, TRUE, "fdr")
mSet<-PlotVolcano(mSet, "volcano_2_",1, "png", 72, width=NA)
mSet<-PlotVolcano(mSet, "volcano_2_",1, "pdf", 72, width=NA)
mSet<-PCA.Anal(mSet)
mSet<-PlotPCAPairSummary(mSet, "pca_pair_0_", "png", 72, width=NA, 5)
mSet<-PlotPCAScree(mSet, "pca_scree_0_", "png", 72, width=NA, 5)
mSet<-PlotPCA2DScore(mSet, "pca_score2d_0_", "png", 72, width=NA, 1,2,0.95,0,0)
mSet<-PlotPCALoading(mSet, "pca_loading_0_", "png", 72, width=NA, 1,2);
mSet<-PlotPCABiplot(mSet, "pca_biplot_0_", "png", 72, width=NA, 1,2)
mSet<-PlotPCA3DLoading(mSet, "pca_loading3d_0_", "json", 1,2,3)
mSet<-GetGroupNames(mSet, "null")
mSet<-PlotPCAPairSummary(mSet, "pca_pair_0_", "pdf", 72, width=NA, 5)
mSet<-PlotPCA2DScore(mSet, "pca_score2d_0_", "pdf", 72, width=NA, 1,2,0.95,0,0)
mSet<-PLSR.Anal(mSet, reg=TRUE)
mSet<-PlotPLSPairSummary(mSet, "pls_pair_0_", "png", 72, width=NA, 5)
mSet<-PlotPLS2DScore(mSet, "pls_score2d_0_", "png", 72, width=NA, 1,2,0.95,0,0)
mSet<-PlotPLS3DScoreImg(mSet, "pls_score3d_0_", "png", 72, width=NA, 1,2,3, 40)
mSet<-PlotPLSLoading(mSet, "pls_loading_0_", "png", 72, width=NA, 1, 2);
mSet<-PlotPLS3DLoading(mSet, "pls_loading3d_0_", "json", 1,2,3)
mSet<-PLSDA.CV(mSet, "T",5, "Q2")
mSet<-PlotPLS.Classification(mSet, "pls_cv_0_", "png", 72, width=NA)
mSet<-PlotPLS.Imp(mSet, "pls_imp_0_", "png", 72, width=NA, "vip", "Comp. 1", 15,FALSE)
mSet<-PlotPLSPairSummary(mSet, "pls_pair_0_", "pdf", 72, width=NA, 5)
mSet<-PlotPLS2DScore(mSet, "pls_score2d_0_", "pdf", 72, width=NA, 1,2,0.95,0,0)
mSet<-PlotPLS.Imp(mSet, "pls_imp_0_", "pdf", 72, width=NA, "vip", "Comp. 1", 15,FALSE)
mSet<-PlotPLS.Classification(mSet, "pls_cv_0_", "pdf", 72, width=NA)
mSet<-PlotHeatMap(mSet, "heatmap_10_", "png", 72, width=NA, "norm", "row", "euclidean", "ward.D","bwm", "overview", T, T, NULL, T, F)
mSet<-PlotHeatMap(mSet, "heatmap_11_", "png", 72, width=NA, "norm", "row", "euclidean", "ward.D","bwm", "detail", T, T, NULL, F, F)
mSet<-PlotHeatMap(mSet, "heatmap_11_", "pdf", 72, width=NA, "norm", "row", "euclidean", "ward.D","bwm", "detail", T, T, NULL, F, F)
mSet<-PlotHeatMap(mSet, "heatmap_12_", "png", 72, width=NA, "norm", "row", "euclidean", "ward.D","bwm", "detail", F, F, NULL, F, F)
mSet<-PlotHeatMap(mSet, "heatmap_12_", "pdf", 72, width=NA, "norm", "row", "euclidean", "ward.D","bwm", "detail", F, F, NULL, F, F)
mSet<-PlotSubHeatMap(mSet, "heatmap_13_", "png", 72, width=NA, "norm", "row", "euclidean", "ward.D","bwm", "tanova", 25, "detail", T, T, F, F)
mSet<-PlotSubHeatMap(mSet, "heatmap_14_", "png", 72, width=NA, "norm", "row", "euclidean", "ward.D","bwm", "tanova", 25, "overview", T, T, F, F)
mSet<-PlotSubHeatMap(mSet, "heatmap_14_", "pdf", 72, width=NA, "norm", "row", "euclidean", "ward.D","bwm", "tanova", 25, "overview", T, T, F, F)
mSet<-PlotSubHeatMap(mSet, "heatmap_15_", "png", 72, width=NA, "norm", "row", "euclidean", "ward.D","bwm", "vip", 15, "overview", T, T, F, F)
mSet<-PlotSubHeatMap(mSet, "heatmap_16_", "png", 72, width=NA, "norm", "row", "euclidean", "ward.D","bwm", "vip", 15, "overview", T, T, F, T)
mSet<-PlotSubHeatMap(mSet, "heatmap_16_", "pdf", 72, width=NA, "norm", "row", "euclidean", "ward.D","bwm", "vip", 15, "overview", T, T, F, T)

#Ctrl vs COVID-19 negatives
feature.nm.vec <- c("")
smpl.nm.vec <- c("")
grp.nm.vec <- c("3")
mSet<-UpdateData(mSet)
mSet<-PreparePrenormData(mSet)
mSet<-Normalization(mSet, "NULL", "LogNorm", "NULL", ratio=FALSE, ratioNum=20)
mSet<-PlotNormSummary(mSet, "norm_3_", "png", 72, width=NA)
mSet<-PlotSampleNormSummary(mSet, "snorm_3_", "png", 72, width=NA)
mSet<-FC.Anal(mSet, 2.0, 0, FALSE)
mSet<-PlotFC(mSet, "fc_3_", "png", 72, width=NA)
mSet<-FC.Anal(mSet, 1.0, 1, FALSE)
mSet<-PlotFC(mSet, "fc_4_", "png", 72, width=NA)
mSet<-Ttests.Anal(mSet, F, 0.05, FALSE, TRUE, "fdr", FALSE)
mSet<-PlotTT(mSet, "tt_2_", "png", 72, width=NA)
mSet<-Ttests.Anal(mSet, T, 1.0, FALSE, TRUE, "fdr", FALSE)
mSet<-PlotTT(mSet, "tt_3_", "png", 72, width=NA)
mSet<-Volcano.Anal(mSet, FALSE, 2.0, 0, F, 0.1, TRUE, "raw")
mSet<-PlotVolcano(mSet, "volcano_2_",1, "png", 72, width=NA)
mSet<-Volcano.Anal(mSet, FALSE, 1.5, 1, T, 0.05, TRUE, "fdr")
mSet<-PlotVolcano(mSet, "volcano_3_",1, "png", 72, width=NA)
mSet<-PlotVolcano(mSet, "volcano_3_",1, "pdf", 72, width=NA)
mSet<-PCA.Anal(mSet)
mSet<-PlotPCAPairSummary(mSet, "pca_pair_0_", "png", 72, width=NA, 5)
mSet<-PlotPCAScree(mSet, "pca_scree_0_", "png", 72, width=NA, 5)
mSet<-PlotPCA2DScore(mSet, "pca_score2d_0_", "png", 72, width=NA, 1,2,0.95,0,0)
mSet<-PlotPCALoading(mSet, "pca_loading_0_", "png", 72, width=NA, 1,2);
mSet<-PlotPCABiplot(mSet, "pca_biplot_0_", "png", 72, width=NA, 1,2)
mSet<-PlotPCA3DLoading(mSet, "pca_loading3d_0_", "json", 1,2,3)
mSet<-GetGroupNames(mSet, "null")
mSet<-PlotPCAPairSummary(mSet, "pca_pair_0_", "pdf", 72, width=NA, 5)
mSet<-PlotPCA2DScore(mSet, "pca_score2d_0_", "pdf", 72, width=NA, 1,2,0.95,0,0)
mSet<-PLSR.Anal(mSet, reg=TRUE)
mSet<-PlotPLSPairSummary(mSet, "pls_pair_0_", "png", 72, width=NA, 5)
mSet<-PlotPLS2DScore(mSet, "pls_score2d_0_", "png", 72, width=NA, 1,2,0.95,0,0)
mSet<-PlotPLS3DScoreImg(mSet, "pls_score3d_0_", "png", 72, width=NA, 1,2,3, 40)
mSet<-PlotPLSLoading(mSet, "pls_loading_0_", "png", 72, width=NA, 1, 2);
mSet<-PlotPLS3DLoading(mSet, "pls_loading3d_0_", "json", 1,2,3)
mSet<-PLSDA.CV(mSet, "T",5, "Q2")
mSet<-PlotPLS.Classification(mSet, "pls_cv_0_", "png", 72, width=NA)
mSet<-PlotPLS.Imp(mSet, "pls_imp_0_", "png", 72, width=NA, "vip", "Comp. 1", 15,FALSE)
mSet<-PlotPLSPairSummary(mSet, "pls_pair_0_", "pdf", 72, width=NA, 5)
mSet<-PlotPLS2DScore(mSet, "pls_score2d_0_", "pdf", 72, width=NA, 1,2,0.95,0,0)
mSet<-PlotPLS.Imp(mSet, "pls_imp_0_", "pdf", 72, width=NA, "vip", "Comp. 1", 15,FALSE)
mSet<-PlotPLS.Classification(mSet, "pls_cv_0_", "pdf", 72, width=NA)
mSet<-PlotHeatMap(mSet, "heatmap_16_", "png", 72, width=NA, "norm", "row", "euclidean", "ward.D","bwm", "overview", T, T, NULL, T, F)
mSet<-PlotHeatMap(mSet, "heatmap_17_", "png", 72, width=NA, "norm", "row", "euclidean", "ward.D","bwm", "detail", T, T, NULL, F, F)
mSet<-PlotHeatMap(mSet, "heatmap_17_", "pdf", 72, width=NA, "norm", "row", "euclidean", "ward.D","bwm", "detail", T, T, NULL, F, F)
mSet<-PlotHeatMap(mSet, "heatmap_18_", "png", 72, width=NA, "norm", "row", "euclidean", "ward.D","bwm", "detail", F, F, NULL, F, F)
mSet<-PlotHeatMap(mSet, "heatmap_18_", "pdf", 72, width=NA, "norm", "row", "euclidean", "ward.D","bwm", "detail", F, F, NULL, F, F)
mSet<-PlotSubHeatMap(mSet, "heatmap_19_", "png", 72, width=NA, "norm", "row", "euclidean", "ward.D","bwm", "tanova", 25, "overview", T, T, F, F)
mSet<-PlotSubHeatMap(mSet, "heatmap_19_", "pdf", 72, width=NA, "norm", "row", "euclidean", "ward.D","bwm", "tanova", 25, "overview", T, T, F, F)
mSet<-PlotSubHeatMap(mSet, "heatmap_20_", "png", 72, width=NA, "norm", "row", "euclidean", "ward.D","bwm", "vip", 15, "overview", T, T, F, T)
mSet<-PlotSubHeatMap(mSet, "heatmap_20_", "pdf", 72, width=NA, "norm", "row", "euclidean", "ward.D","bwm", "vip", 15, "overview", T, T, F, T)

\# Comorbidities
\## Heatmaps: were obtained as other heatmaps using MetaboAnalystR.
\## K-means: example with McCabe classification, the procedure was the same for Charlson index.
mSet<-InitDataObjects("conc", "stat", FALSE)
mSet<-Read.TextData(mSet, "COVID_Lipidomics_McCabe.csv", "rowu", "disc");
mSet<-SanityCheckData(mSet)
mSet<-SanityCheckData(mSet)
mSet<-RemoveMissingPercent(mSet, percent=0.5)
mSet<-ImputeMissingVar(mSet, method="knn_var")
mSet<-SanityCheckData(mSet)
mSet<-FilterVariable(mSet, "none", "F", 25)
mSet<-PreparePrenormData(mSet)
mSet<-Normalization(mSet, "NULL", "LogNorm", "NULL", ratio=FALSE, ratioNum=20)
mSet<-PlotNormSummary(mSet, "norm_0_", "png", 72, width=NA)
mSet<-PlotSampleNormSummary(mSet, "snorm_0_", "png", 72, width=NA)
mSet<-Kmeans.Anal(mSet, 3)
mSet<-PlotKmeans(mSet, "km_0_", "png", 72, width=NA, "default", "F")
mSet<-PlotClustPCA(mSet, "km_pca_0_", "png", 72, width=NA, "default", "km", "F")
mSet<-Kmeans.Anal(mSet, 3)
mSet<-PlotKmeans(mSet, "km_1_", "png", 72, width=NA, "colblind", "F")
mSet<-PlotClustPCA(mSet, "km_pca_1_", "png", 72, width=NA, "colblind", "km", "F")
mSet<-PlotClustPCA(mSet, "km_pca_1_", "pdf", 72, width=NA, "colblind", "km", "F")

**Biomarker analysis:**

The following piece of code was used to calculate the MCCV model and to plot the ROC curves to compare COVID-19-negative patients and COVID-19-positive patients. We followed the same procedure for the other comparisons.

library(MetaboAnalystR)
mSet<-InitDataObjects("conc", "roc", FALSE);
mSet<-Read.TextData(mSet, "COVID_Lipidomics_COVIDneg-vs-COVIDpos.csv", "rowu", "disc");
mSet<-SanityCheckData(mSet)
mSet<-ContainMissing(mSet)
mSet<-ContainsMetaDataFile(mSet)
mSet<-SanityCheckData(mSet)
mSet<-ContainMissing(mSet)
mSet<-ContainsMetaDataFile(mSet)
mSet<-RemoveMissingPercent(mSet, percent=0.5)
mSet<-ImputeMissingVar(mSet, method="knn_var")
mSet<-SanityCheckData(mSet)
mSet<-ContainMissing(mSet)
mSet<-ContainsMetaDataFile(mSet)
mSet<-FilterVariable(mSet, "none", "F", 25)
mSet<-PreparePrenormData(mSet)
mSet<-Normalization(mSet, "NULL", "LogNorm", "NULL", ratio=TRUE, ratioNum=20)
mSet<-PlotNormSummary(mSet, "norm_0_", "png", 72, width=NA)
mSet<-PlotSampleNormSummary(mSet, "snorm_0_", "png", 72, width=NA)
mSet<-SetAnalysisMode(mSet, "explore")
mSet<-PrepareROCData(mSet)
mSet<-PerformCV.explore(mSet, "svm", "svm", 2)
mSet<-PlotProbView(mSet, "cls_prob_0_", "png", 72, -1, 0, 0)
mSet<-PlotImpVars(mSet, "cls_imp_0_", "png", 72, -1, "freq", 15);
mSet<-PlotAccuracy(mSet, "cls_accu_0_", "png", 72)
mSet<-PlotROC(mSet, "cls_roc_0_", "png", 72, 0, "threshold", 0, 0, "fpr", 0.5)
mSet<-PlotROC(mSet, "cls_roc_0_", "pdf", 72, 0, "threshold", 0, 0, "fpr", 0.5)
mSet<-PlotROC(mSet, "cls_roc_1_", "png", 72, 6, "threshold", 1, 0, "fpr", 0.2)
mSet<-PlotROC(mSet, "cls_roc_1_", "pdf", 72, 6, "threshold", 1, 0, "fpr", 0.2)
mSet<-PlotProbView(mSet, "cls_prob_1_", "png", 72, 6, 0, 0)
mSet<-PlotProbView(mSet, "cls_prob_1_", "pdf", 72, 6, 0, 0)
mSet<-PlotImpVars(mSet, "cls_imp_1_", "png", 72, 6, "freq", 15);
mSet<-PlotImpVars(mSet, "cls_imp_1_", "pdf", 72, 6, "freq", 15);
mSet<-PlotImpVars(mSet, "cls_imp_2_", "png", 72, 6, "imp", 15);
mSet<-PlotImpVars(mSet, "cls_imp_2_", "pdf", 72, 6, "imp", 15);
mSet<-SetAnalysisMode(mSet, "test") mSet<-PrepareROCData(mSet)
mSet<-CalculateFeatureRanking(mSet)
selected.cmpds <- c("CAR16_1/FA22_6O");
selected.smpls <- c()
mSet<-SetCustomData(mSet, selected.cmpds, selected.smpls)
mSet<-PerformCV.test(mSet, "svm", 2)
mSet<-PlotROC(mSet, "cls_test_roc_0_", "pdf", 72, 0, "threshold", 0, 0, "fpr", 0.5)
mSet<-PlotProbView(mSet, "cls_test_prob_0_", "pdf", 72, -1, 0, 0)
mSet<-PlotTestAccuracy(mSet, "cls_test_accu_0_", "png", 72)
mSet<-GetAccuracyInfo(mSet)
mSet<-PlotROC(mSet, "cls_test_roc_1_", "pdf", 72, 0, "threshold", 1, 0, "fpr", 0.2)

**Enrichment analysis:**

library(MetaboAnalystR)
#Name conversion
mSet<-InitDataObjects("NA", "utils", FALSE)
cmpd.vec<-c("CE(16:0)","CE(16:1)","CE(17:1)","CE(18:0)","CE(18:1)","CE(18:2)","CE(18:3)","CE(20:2)","CE(20:3)","CE(20:4)","CE(20:5)","CE(22:4)","CE(22:5)","CE(22:6)","DG(34:1)","DG(34:2)","DG(34:3)","DG(36:0)","DG(36:1)","DG(36:2)","DG(36:3)","DG(36:4)","DG(40:4)","lauric acid","Myristic acid","Palmitic acid","palmitoleic acid","palmitelaidic acid","Stearic acid","Oleic acid","epoxy-stearic acid","12,13-DiHOME(9)","9,10-DiHOME(12)","hydroxy-epoxy-stearic acid","9,12,13-TriHOME(10)","Linoleic acid","9-HODE","DiHODE","9,10-EpOME(12)","12,13-EpOME(9)","Linolenic acid (iso1)","Linolenic acid (iso2)","15,16-epoxy-13-OH-9Z,11E-octadecadienoic acid","OxoODE (iso1)","OxoODE (iso2)","Stearidonic acid (iso1)","Nonadecenoic acid","Eicosenoic acid","11,13-Eicosadienoic acid","bishomo-y-linolenic acid (iso1)","bishomo-y-linolenic acid (iso2)","bishomo-y-linolenic acid (iso3)","15-HETrE","Arachidonic acid","12-HETE","15-HETE","20-HETE","HETE (iso1)","eicosapentaenoic acid","HpEPE","Adrenic acid","omega3-docosapentaenoic acid","omega6-docosapentaenoic acid","Docosahexaenoic acid","17-HDHA","PC(14:0/0:0)","PC(0:0/14:0)","LPC(15:0)","PC(15:0/0:0)","PC(0:0/15:0)","LPC(16:0)","LPC(O-16:0)","PC(16:0/0:0)","PC(0:0/16:0)","LPC(O-16:1)","PC(16:1/0:0)","PC(0:0/16:1)","PC(17:0/0:0)","PC(0:0/17:0)","PC(17:1/0:0)","PC(0:0/17:1)","LPC(18:0)","LPC(O-18:0)","PC(18:0/0:0)","PC(0:0/18:0)","LPC(18:1)","PC(18:1/0:0)","PC(0:0/18:1)","LPC(18:2)","PC(18:2/0:0)","PC(0:0/18:2)","PC(18:3/0:0)","PC(0:0/18:3)","PC(19:0/0:0)","PC(0:0/19:0)","LPC(20:0)","PC(20:0/0:0)","PC(0:0/20:0)","PC(20:1/0:0)","PC(0:0/20:1)","LPC(20:2)","PC(20:2/0:0)","PC(0:0/20:2)","LPC(20:3)","PC(20:3/0:0)","PC(0:0/20:3)","PC(20:4/0:0)","PC(0:0/20:4)","PC(20:5/0:0)","PC(0:0/20:5)","PC(22:3/0:0)","PC(0:0/22:3)","PC(22:4/0:0)","PC(0:0/22:4)","PC(22:5/0:0)","PC(0:0/22:5)","PC(22:6/0:0)","PC(0:0/22:6)","PE(14:0/0:0)","PE(0:0/14:0)","PE(15:0/0:0)","PE(0:0/15:0)","PE(16:0/0:0)","PE(0:0/16:0)","PE(16:1/0:0)","PE(0:0/16:1)","PE(17:0/0:0)","PE(17:1/0:0)","PE(0:0/17:1)","PE(18:0/0:0)","PE(0:0/18:0)","PE(18:1/0:0)","PE(0:0/18:1)","PE(18:2/0:0)","PE(0:0/18:2)","PE(18:3/0:0)","PE(0:0/18:3)","PE(20:1/0:0)","PE(0:0/20:1)","PE(20:2/0:0)","PE(20:3/0:0)","PE(0:0/20:3)","PE(20:4/0:0)","PE(0:0/20:4)","PE(20:5/0:0)","PE(0:0/20:5)","PE(22:4/0:0)","PE(0:0/22:4)","PE(22:5/0:0)","PE(22:6/0:0)","PE(0:0/22:6)","N-palmitoyl ethanolamine","N-stearoyl-ethanolamine","N-oleoyl ethanolamine","N-Linoleoyl Ethanolamine","PC(30:0)","PC(31:0)","PC(32:0)","PC(32:1)","PC(32:2)","PC(33:0)","PC(33:1)","PC(33:2)","PC(34:0)","PC(34:1)","PC(34:2)","PC(34:3)","PC(34:4)","PC(35:1)","PC(35:2)","PC(35:4)","PC(36:1)","PC(36:2)","PC(36:3)","PC(36:4)","PC(36:5)","PC(38:2)","PC(38:3)","PC(38:4)","PC(38:5)","PC(38:6)","PC(40:4)","PC(40:5)","SM(32:1)","SM(32:2)","SM(33:1)","SM(34:1)","SM(34:2)","SM(35:1)","SM(36:0)","SM(36:1)","SM(36:2)","SM(38:1)","SM(38:2)","SM(39:1)","SM(40:1)","SM(40:2)","SM(41:1)","SM(41:2)","SM(42:1)","SM(42:2)","SM(42:3)","SM(43:1)","SM(43:2)","Sphinganine-1-P","Sphingosine-1-P","Dehydroepiandrosterone","Cortisol","Cortisone","Taurolithocholic acid","Deoxycholic acid","Chenodeoxycholic Acid","Glycoursodeoxycholic acid","Glycochenodeoxycholic acid","Glycodeoxycholic acid","Deoxycholic acid (iso1)","Deoxycholic acid-iso2","Ursodeoxycholic acid","Cholic acid","Glycocholic Acid","Taurocholic acid","TG(46:0)","TG(46:1)","TG(46:2)","TG(47:0)","TG(47:1)","TG(48:0)","TG(48:1)","TG(48:2)","TG(48:3)","TG(50:0)","TG(50:1)","TG(50:2)","TG(50:3)","TG(50:4)","TG(51:2)","TG(51:3)","TG(52:1)","TG(52:2)","TG(52:3)","TG(52:4)","TG(52:5)","TG(52:6)","TG(54:2)","TG(54:3)","TG(54:4)","TG(54:5)","TG(54:6)","TG(54:7)","TG(56:5)","TG(56:6)","TG(56:7)","TG(58:8)")
mSet<-Setup.MapData(mSet, cmpd.vec)
mSet<-CrossReferencingAPI(mSet, "name")
mSet<-CreateMappingResultTable(mSet)
#With the obtained datatable, we matched the compound names with their respective HMDB, KEGGS, PubChem, ChEBI, and MetLin identifications. As the most completed identification were the PubChem ones, we created a database with PubChem identifications instead of compound names with each lipid concentrations.

#All three comparisons (Control versus COVID+; Control versus COVID-; and COVID+ versus COVID-) were performed using the following:
mSet<-InitDataObjects("conc", "msetqea", FALSE)
mSet<-Read.TextData(mSet, "COVID_Lipidomics_enrichment_Ctrl_COVIDpos.csv", "rowu", "disc"); #The file name was changed in each comparison
mSet<-SanityCheckData(mSet)
mSet<-ContainMissing(mSet)
mSet<-SanityCheckData(mSet)
mSet<-ContainMissing(mSet)
mSet<-RemoveMissingPercent(mSet, percent=0.5)
mSet<-ImputeMissingVar(mSet, method="knn_var")
mSet<-CrossReferencing(mSet, "pubchem", lipid = T);
mSet<-CreateMappingResultTable(mSet)
mSet<-GetCandidateList(mSet);
mSet<-PreparePrenormData(mSet)
mSet<-Normalization(mSet, "NULL", "LogNorm", "NULL", ratio=FALSE, ratioNum=20)
mSet<-SetMetabolomeFilter(mSet, F);
mSet<-SetCurrentMsetLib(mSet, "smpdb_pathway", 2);
mSet<-CalculateGlobalTestScore(mSet)
mSet<-PlotQEA.Overview(mSet, "qea_0_", "net", "png", 72, width=NA)
mSet<-PlotEnrichDotPlot(mSet, "qea", "qea_dot_0_", "png", 72, width=NA)
mSet<-PlotQEA.Overview(mSet, "qea_0_", "net", "pdf", 72, width=NA)
mSet<-PlotEnrichDotPlot(mSet, "qea", "qea_dot_0_", "pdf", 72, width=NA)

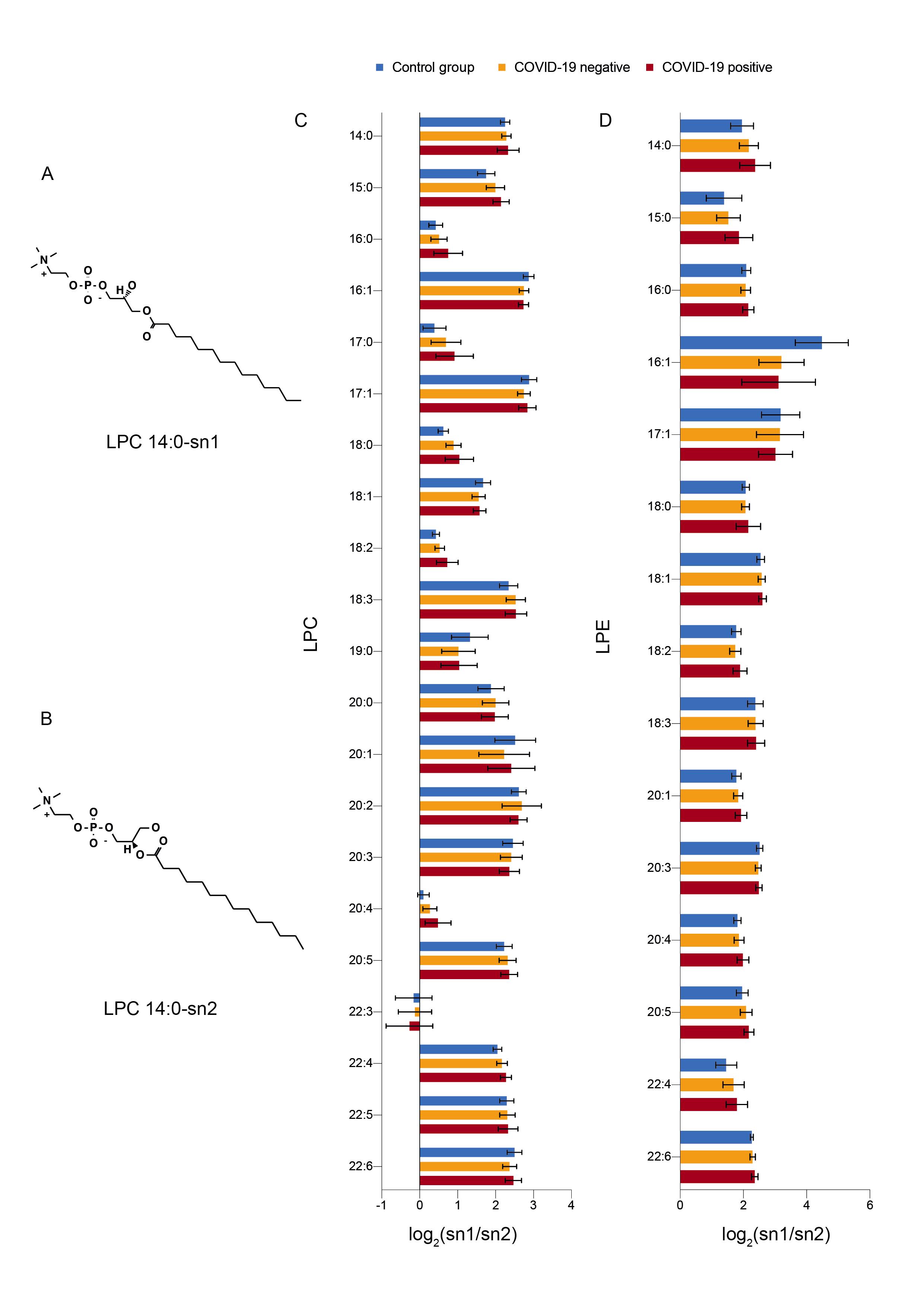

**Supplementary Figure 1.** Examples of a lysophosphatidylcholine (LPC) with the acyl chain (14:0) attached to (A) sn1 position and (B) sn2 position. Fold change of sn1 and sn2 concentration of (C) lysophosphatidylcholines (LPC) and (D) lysophosphatidylethanolamines (LPE) in the three studied groups. We did not find any differences in the proportion of sn1/sn2.

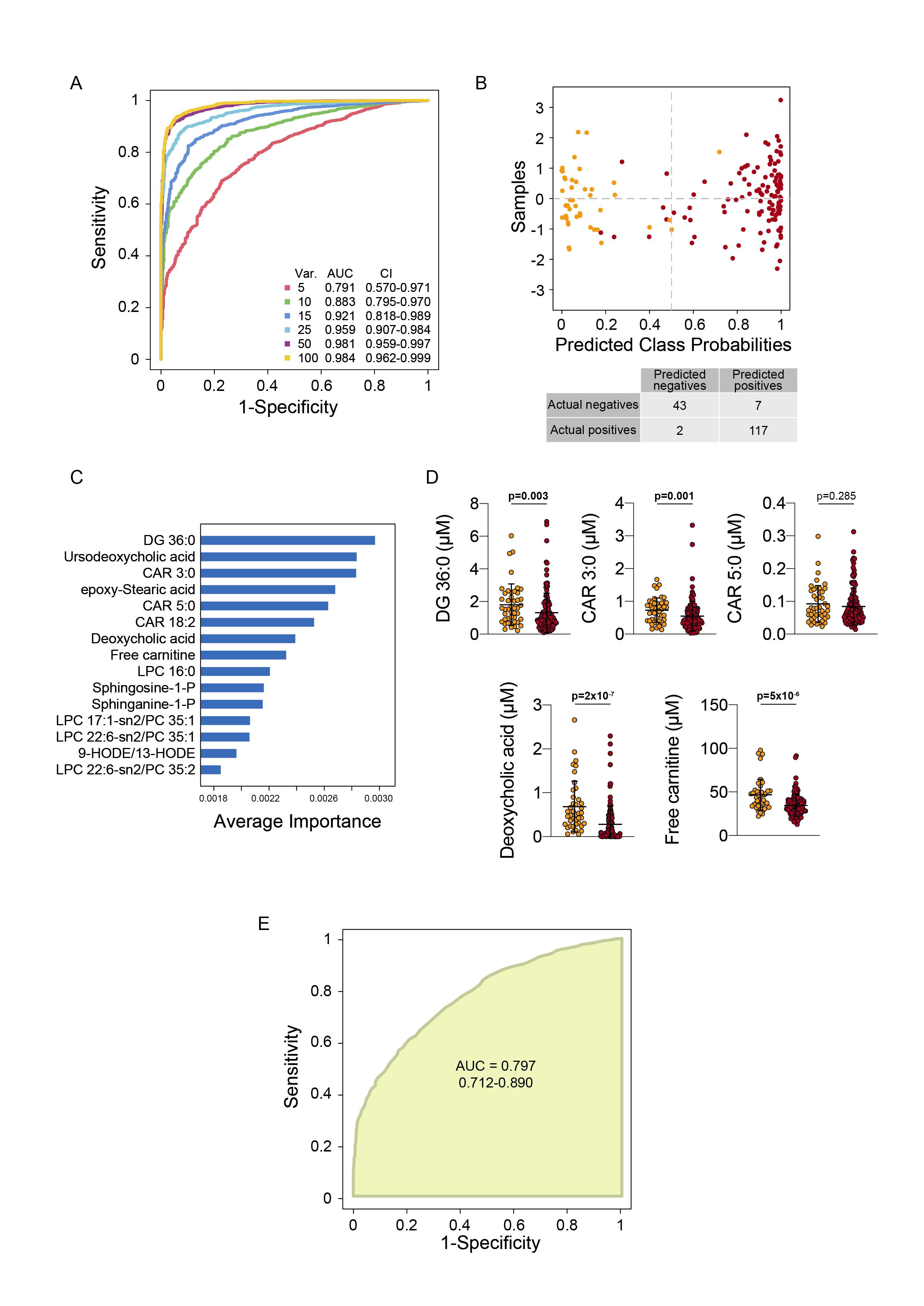

**Supplementary Figure 2.** Receiver operating characteristic (ROC) curves (a) and confusion matrix (B) of Monte Carlo cross validation model to test the discriminatory ability of the lipidomic profile. Top 15 variables classified as the most important by the model (C). Selected lipid concentrations in serum of COVID-19-negative patients (yellow dots) and COVID-19-positive patients (red dots). (E) ROC curve to discriminate between COVID-19-positive and COVID-19-negative patients.
